# Effect of age, sex and BMI on resting ECG intervals and their variabilities in healthy adults

**DOI:** 10.64898/2026.03.07.26347862

**Authors:** Qihou Zhou

## Abstract

**Objective:** While there are numerous reports on heart rate and its variabilities, a detailed analysis of the component intervals for healthy adults in well controlled condition is lacking. This study analyzes the effect of age, sex, and Body Mass Index (BMI) on nine resting electrocardiogram (ECG) intervals and their intra-individual variabilities in healthy adults under the same testing environment.

**Methods:** Using the “Autonomic Aging” dataset, ECG recordings from 1,121 healthy volunteers (ages 18–92) were processed. The study employed a specialized segmentation algorithm to identify key ECG markers. We analyze statistically how age, BMI, and sex impact the durations and variabilities of nine ECG intervals.

**Results:** Fifty years of age serves as a critical transition age for cardiac aging for all subjects as a whole. Above this age, the active interval, which is the combined atrial and ventricular conduction time, increases three times faster than at a younger age, primarily driven by lengthening of depolarization times. Compared to the opposite sex, older low-BMI males have a longer atrial conduction time, and older low-BMI females have a larger variability in the ventricular conduction time. High BMI increases the heart rate by reducing the length of the idle interval, i.e., the isoelectric segment at the end of a cardiac cycle. The rate increase is more pronounced among older subjects than younger ones. High BMI males start to exhibit an elevated heart rate and larger variability in the atrial conduction time in their 30s. High BMI females start to show a larger variability in the ventricular repolarization time around 50 years old.

**Conclusion:** Age, BMI, and sex all have major impacts on the ECG intervals and their variability. A resting heart behaves largely like a pulse width modulation system, with a stable active interval and an adjustable idle interval to meet the varying needs for cardiac output. The durations and variabilities of the active interval, more than those of the RR interval, are indicators of a heart’s health condition. A young and healthy heart tends to have a shorter duration and smaller variability in the active interval.

## 1. INTRODUCTION

The measurement and analysis of electrocardiographic (ECG) intervals, such as the PR interval and QRS duration, are essential for assessing cardiac health and autonomic function. In healthy individuals, these intervals are dynamically regulated by the autonomic nervous system. Increased sympathetic stimulation during physical exertion or sympathomimetic infusions enhances atrioventricular (AV) nodal conduction and shortens cycle length, whereas parasympathetic conduction predominates at rest and modulates the beat-to-beat variability of RR and PR intervals (Fisher et al. 2015; Rosenberg et al., 2020). However, accumulating data indicate that age, sex, and body mass index (BMI) have distinct and interacting signatures on ECG intervals and their variabilities (Giovanardi et al. 2022; Ahmadi et al. 2023; Kumari et al., 2023). Evidence underscores that these parameters are not static but vary systematically across demographic strata, with significant implications for risk management and emerging AI-based phenotyping (Giovannardi et al., 2022; Hempel et al., 2025).

Age exerts a dominant influence on ECG traits and autonomic markers, particularly in later adulthood. Large population studies demonstrate strong age associations for the PR interval, QT/QTc, QRS duration, and the frontal QRS–T angle, with an increase in PR, QTc, and leftward QRS axis shift and higher prevalence of ECG abnormalities as age advances (Macfarlane et al., 1994; Tan et al., 2016; Cavarretta et al., 2022; Giovannardi et al., 2022; Ahmadi et al., 2023). AI models trained to estimate “ECG age” show that P wave duration, PR interval, QRS duration, and QTc all increase with age, and such increases are associated with elevated risks of atrial fibrillation, heart failure, and mortality (Hempel et al. 2022). Long term cohort data further indicate that both short and long PR intervals, as well as temporal changes in PR, are associated with higher rates of atrial fibrillation, heart failure, ventricular arrhythmias, syncope, and all-cause mortality, emphasizing the need to distinguish benign age-related variation from truly high-risk conduction phenotypes (Zeng et al., 2025).

Sex differences remain evident across the adult lifespan, leaving distinct signatures on ECG. Women generally have higher resting heart rates and longer QT or QTc intervals, whereas men exhibit longer PR and QRS intervals, and higher QRS and ST amplitudes (Carbone et al., 2020; Ahmadi et al., 2023; Kittnar, 2023; Ngoc et al., 2025). Experimental and clinical work indicates that women’s longer QT and reduced repolarization reserve, modulated by sex hormones, contribute to a higher susceptibility to drug-induced and congenital torsades de pointes compared with men (Abi-Gerges, 2004; Carbone et al., 2020; Prajapati et al., 2022). These sex specific repolarization patterns and their evolution with age underscore the need for ECG interpretation criteria in older adults that explicitly incorporate sex, rather than extrapolating from middle-aged or male-derived norms (Carbone et al., 2020; Giovanardi et al., 2022; Ahmadi et al., 2023; Kittnar, 2023).

Elevated Body Mass Index (BMI) significantly alters the heart’s structure and function, increasing the cardiovascular system’s workload and the risk of chronic disease. Higher BMI is causally associated with prolonged P-wave duration and QTc interval, which are markers of atrial and ventricular arrhythmogenic risk, as shown by Mendelian randomization studies (Ardissino et al., 2023; Yang et al., 2024). Increased BMI also correlates with lengthened conduction times, such as PR interval and QRS duration, with sex differences noted—females tend to have narrower QRS complexes than males at similar BMI levels (Ma et al., 2022; Mrad et al., 2021; Rao et al., 2021). Obesity phenotypes influence ECG abnormalities, including left atrial enlargement and inferior T-wave inversions, particularly in physically active young males, while the relationship with left ventricular hypertrophy varies by ECG criteria (Lin et al., 2021). Advanced AI models can predict BMI from ECG signals, introducing a novel biomarker (delta-BMI) that stratifies cardiometabolic risk beyond measured BMI (Pastika et al., 2024). Additionally, obesity is associated with prolonged repolarization markers like the *T*_*peak*_ − *T*_*end*_ interval and increased P-wave dispersion, which may contribute to arrhythmia susceptibility (Dykiert et al., 2024; Partika et al., 2024). The increase in P wave duration and dispersion from normal weight to obesity is consistent with progressive atrial electrical remodeling that may presage atrial fibrillation (Tobenna, 2021; Kumari et al., 2023; Yang et al., 2024).

Despite this progress, further work is needed to explicitly stratify healthy cohorts by sex, age, and BMI, especially with long enough ECG recordings to establish the variabilities as well. Many reference datasets do not have all the age, sex, and BMI information and often under-represent obese or older individuals, which obscures cross-interaction effects and limits generalizability to high-BMI and aging populations (Giovanardi et al., 2022; Jiang et al., 2024). Previous studies often focus on the *RR* interval and do not contain the intra-individual variabilities of other intervals. This study addresses these gaps using the “Autonomic Aging” database of 1,121 subjects (ages 18–92) measured under uniform resting conditions, employing a refined segmentation algorithm that computes nine interval lengths and their variabilities. The analysis here emphasizes the “active interval”, i.e., the total atrial and ventricular conduction time, and the “idle interval”, i.e., the isoelectric segment before atrial depolarization. By performing a systematic analysis of nine ECG interval durations and their variabilities, this research aims to provide a baseline for the “normality map” and the distinct features of each stratification of age, BMI, and sex.

## 2. DATABASE, INTERVAL DEFINITIONS, AND ECG SEGMENTATION

The “Autonomic Aging” dataset, provided by Andy Schumann and Karl-Jürgen Bär (2021, 2022), contains high-resolution biological signals designed to study the impact of healthy aging on cardiovascular regulation. It is available for public access at the Physionet repository (Goldberger et al., 2000). This database comprises Lead II electrocardiogram (ECG) recordings from 1,121 healthy volunteers ranging in age from 18 to 92 years, all collected at Jena University Hospital, Germany. Signals were sampled at 1000 Hz in a controlled environment at 22°C and are accompanied by demographic metadata, including age group, sex, and body mass index. The data were processed for this study using a 0.2 Hz high-pass filter followed by a 20-sample running average to eliminate the 50 Hz power-line interference and its harmonics. Although most records exceed 15 minutes in length, only the final 10 minutes from each subject were analyzed to maintain consistency and minimize potential bias.

In this study, we use *P, R*, and *T* to denote the peak locations of the P-wave, QRS complex, and the T-wave, respectively, while *Q* and *S* indicate the two local minima of the QRS complex on either side of *R*. We designate the beginning of the P-wave as *P*_*b*_ and the end of the T-wave as *T*_*e*_. To reference an interval in general terms, we use the beginning and ending markers, such as *RR* or *P*_*b*_*T*. The time duration between two markers, e.g., *P*_*b*_ and *T*_*e*_, and its intra-individual variability, which is computed from the standard deviation, are denoted as 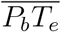 and 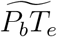_*e*_, respectively. All interval lengths and their variabilities are measured in milliseconds (ms) unless otherwise stated.

Our algorithm begins by identifying all *R* points by analyzing the first- and second-order differences, following the strategies described by Pan and Tompkins (1985). Once the *R* points are located, the segmentation algorithm identifies *Q, S, T*, and *P* points, the end of the T-wave (*T*_*e*_), and the beginning of the P-wave (*P*_*b*_) based on the following general rules: *Q* and *S* points are identified as the first local minima within a distance of 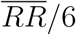 before and after *R*, respectively. The *T* point is defined as the largest local maximum between *S* and 0.6*RR. P* is found as either the second peak or the only local maximum in the interval having a length of 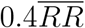 ending at the trailing *Q*. We use the maximum second-order difference between *T* (*P*) and the midpoint between *T* and *P* as *T*_*e*_ (*P*_*b*_). The identified points from hundreds of heartbeats were visually inspected, and erroneous identifications were found to be rare.

Our primary focus is on the durations and variabilities of *RR, P*_*b*_*T*_*e*_, *T*_*e*_*P*_*b*_; *P*_*b*_*Q, P*_*b*_*P, PQ*; *QT*_*e*_, *QS*, and *ST*_*e*_. *P*_*b*_*T*_*e*_, referred to as the “active interval”, represents the entire duration of atrial and ventricular conduction times. *T*_*e*_*P*_*b*_ is the isoelectric segment between two consecutive beats, commonly known as the *TP* interval. *T*_*e*_*P*_*b*_ is referred to as the “idle interval” due to the lack of electrical and mechanical activities. 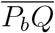 is largely the atrial conduction time and the delay duration at the SA node. It is slightly longer than the traditionally defined *PR* interval, as 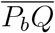 includes the initial part of ventricular depolarization. 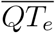 represents the ventricular conduction time duration. Our 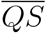 is slightly shorter than the traditional *QRS* duration, and similarly, our 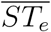 is slightly longer than the traditional *ST* interval. We have excluded traditional durations of *PR, ST*, and *QRS* because the isoelectric segments around the *QRS* complex are often poorly defined in realistic ECG recordings. The definitions defined here result in more accurate statistics, while qualitative comparisons with previous studies remain largely unaffected.

## 3. RESULTS

### 3.1. Age dependence of heart intervals and their variabilities

We exclude records whose 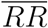 falls outside the 500–1550 ms range or whose 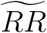 exceeds 150 ms to minimize the impact of outliers, which frequently arise from noisy samples. Given that the determination of *P*_*b*_ is subject to greater measurement uncertainty than the *R* point, records with 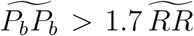 are also excluded. The number of subjects meeting the basic inclusion criteria is 958 out of 1121 total records. The Autonomic Aging dataset defines 15 age groups spanning 18 to 92 years. The age range of the first group is 18-19 years, and that of the last group is 84-92 years. The remaining groups have a 5-year range interval, starting at 20 years for group 2. To reduce statistical fluctuations and ensure a reasonable sample size, we combined the 15 groups into four cohorts for basic age-dependent analyses. The age ranges for the four cohorts are: C1: 15-29 years; C2: 30-44 years; C3: 45-54 years; C4: 55-92 years. The number of samples meeting the inclusion criteria for each cohort is detailed in Table 1, with the numbers in parentheses indicating the male-to-female ratio.

**Table 1.**
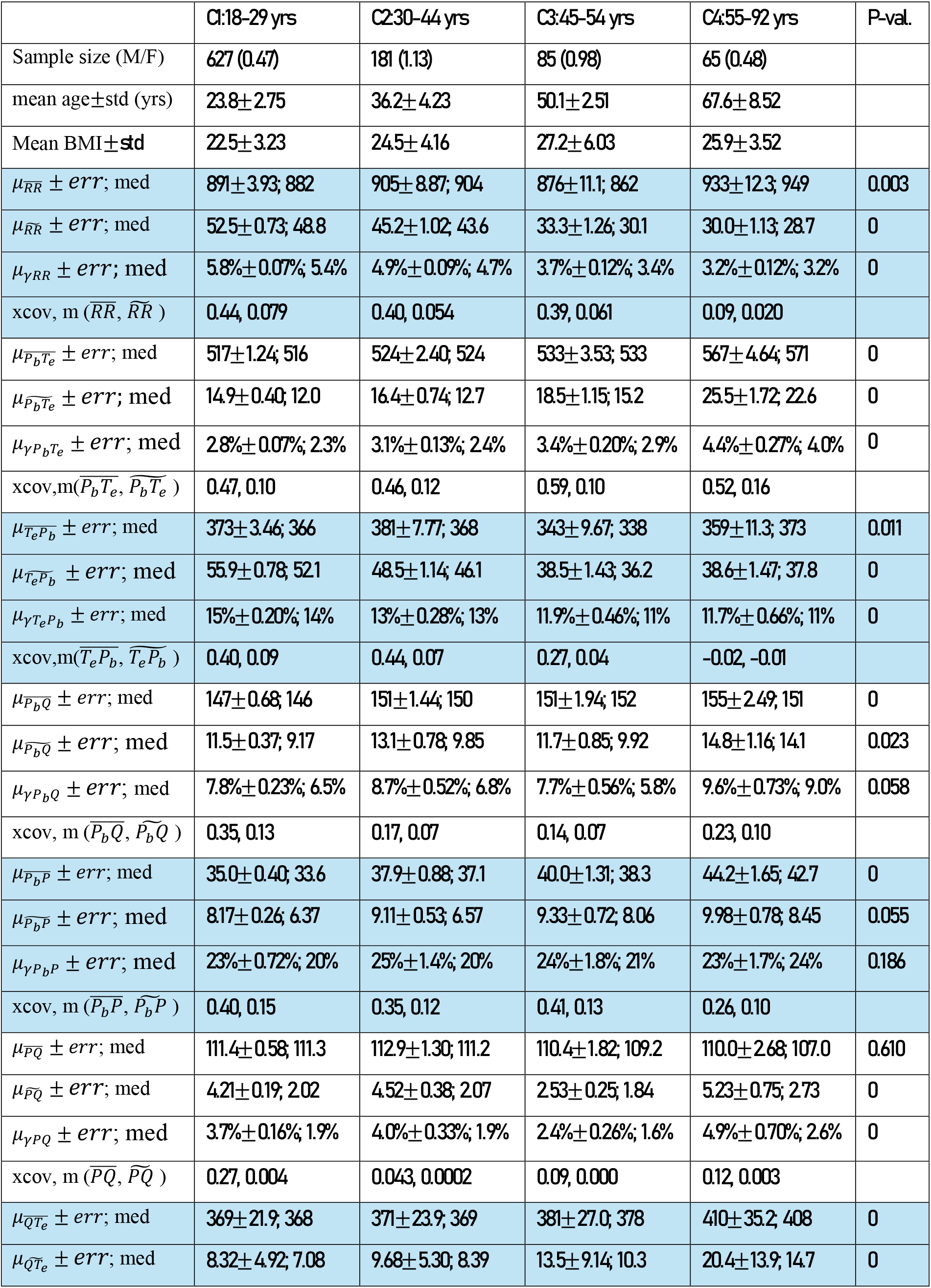
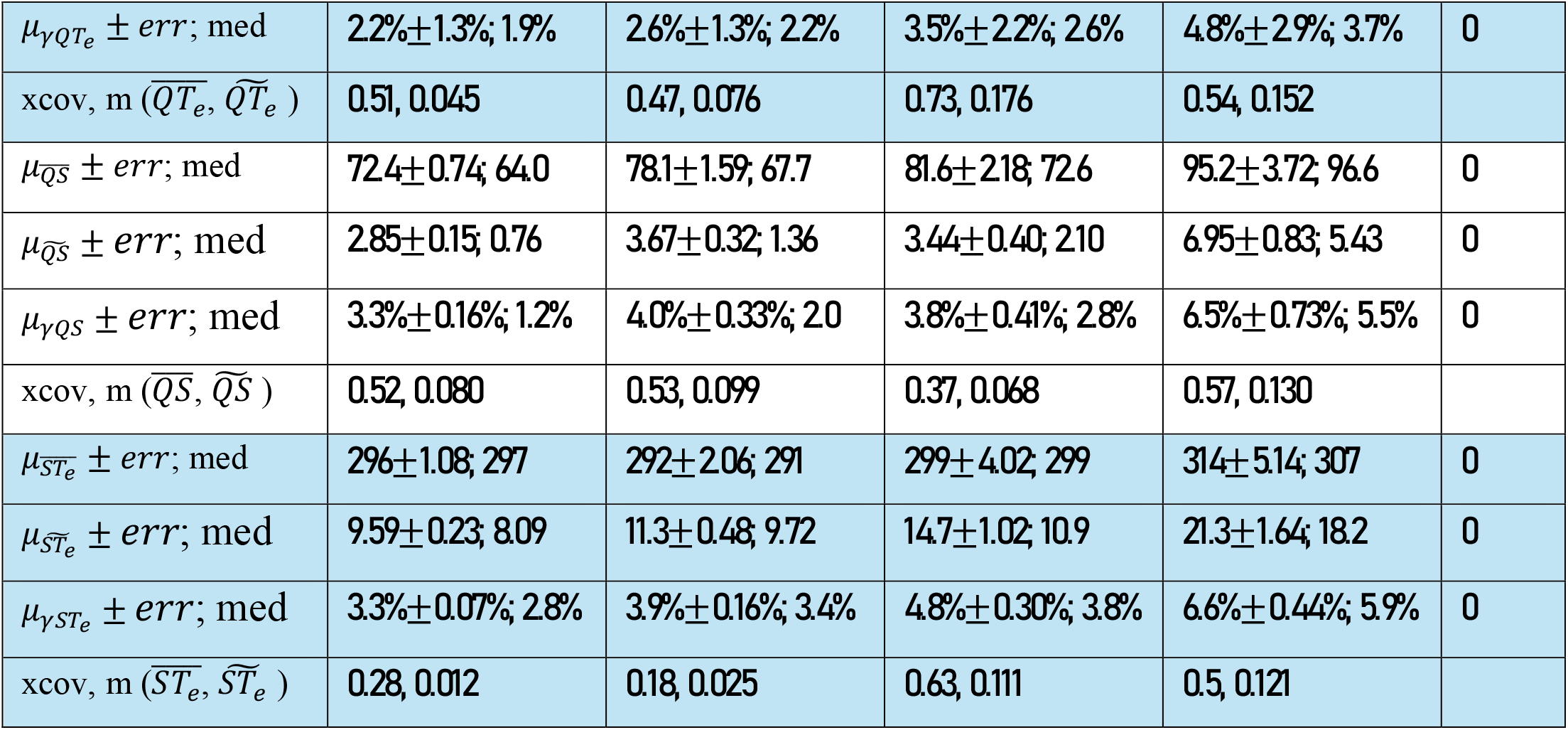
Mean ECG intervals and their variabilities for 4 age cohorts.

The mean ages and standard deviations for each cohort are estimated by adopting the midpoint of each group’s age range as the representative age for all individuals in that group, as more precise ages are not provided in the dataset. For example, all subjects in group 2 (20-24 years) are assumed to be 22.5 years old. In Table 1, we list the means and medians of 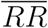 and the eight other intervals, along with their associated variabilities, for the four cohorts. These intervals allow us to examine the components of *RR*, atrial, and ventricular conduction times. In row 4 of Table 1, 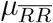 is the mean of 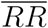 from a cohort using the 5-95 percentile data range. Its error, computed by dividing the standard deviation of individual 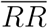 by the square root of the sample size, is listed after the “*±*” sign. The number following the error is the median, which is more robust against outliers but less accurate for normal distributions. In row 6, 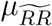 is the mean of 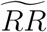, i.e., the mean 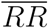 variability within a cohort. To account for the frequent positive correlation between the duration of an interval and its variability, we compute their ratio, *γ*. For instance, *γ*_*RR*_ is the ratio of 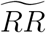 to 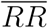 for each person, and 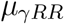, listed in row 7, is the mean *γ*_*RR*_ of a cohort. The errors for 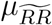 and *µ*_*γRR*_ are calculated in the same manner as that for 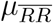. *xcov* and *m* in row 8 denote the cross-correlation coefficient and the slope of the linear fit between the duration of an interval and its variability, respectively. The mean, variability, relative variability, and correlations for other intervals are computed and defined analogously to the 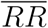 interval. The p-values in the last column evaluate the null hypothesis that all means are equal, using the Welch t-test. A p-value smaller than 5% is commonly considered to indicate that the null hypothesis can be rejected with acceptable significance. A p-value smaller than 0.001 is reported as zero in Table 1. The errors listed in Table 1 can be used to estimate the statistical significance of the difference between any two means. For a normal distribution, if the difference in means exceeds two combined standard deviations (calculated via the square root of the sum of the variances), the corresponding p-value is typically less than 5% for the sample sizes specified in Table 1.

The unweighted mean 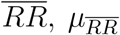, for the four cohorts is 901 ms, which is equivalent to 66.6 beats per minute (bpm). The minimum 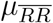 occurs in the 45-54 years old cohort, which exhibits the highest BMI among the four cohorts. A more detailed plot of the median 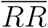 with a finer age resolution is shown in Figure 1(a) along with the median 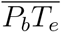 and 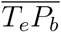. The minimum number of samples is 30 at the mean age of 60.6 yrs. The minimum 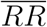 occurring around 50 years is similar to the report by Quer et al. (2020). The mean intra-person variability, 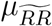, varies significantly across the age cohorts, from 52.5 ms for Cohort 1 to 30 ms for Cohort 4. Intra-person *RR* variability 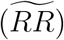 is well-documented to decrease with age. 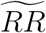, as shown in Figure 1(b), decreases linearly until 45 years. Above 50 years, the median 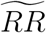 remains largely constant at 30 ms, a characteristic similar to that reported by Quer et al. (2020). The mean ratio of 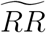 to 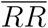 (*µ*_*γRR*_) shows a trend parallel to 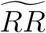, indicating that 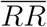 plays only a minor role in the age variation of 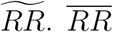 and 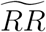 are most strongly correlated in the youngest cohort, with a cross-correlation coefficient of 0.44. We note that Lee and Byun (2023) also analyzed the RR interval length and variability of the Autonomic Aging dataset for age prediction. While the mean 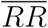 is about the same, our mean 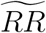 is 20% smaller, which can be due to our more restrictive inclusion criteria and possibly a more robust segmentation algorithm.

**Figure 1.**
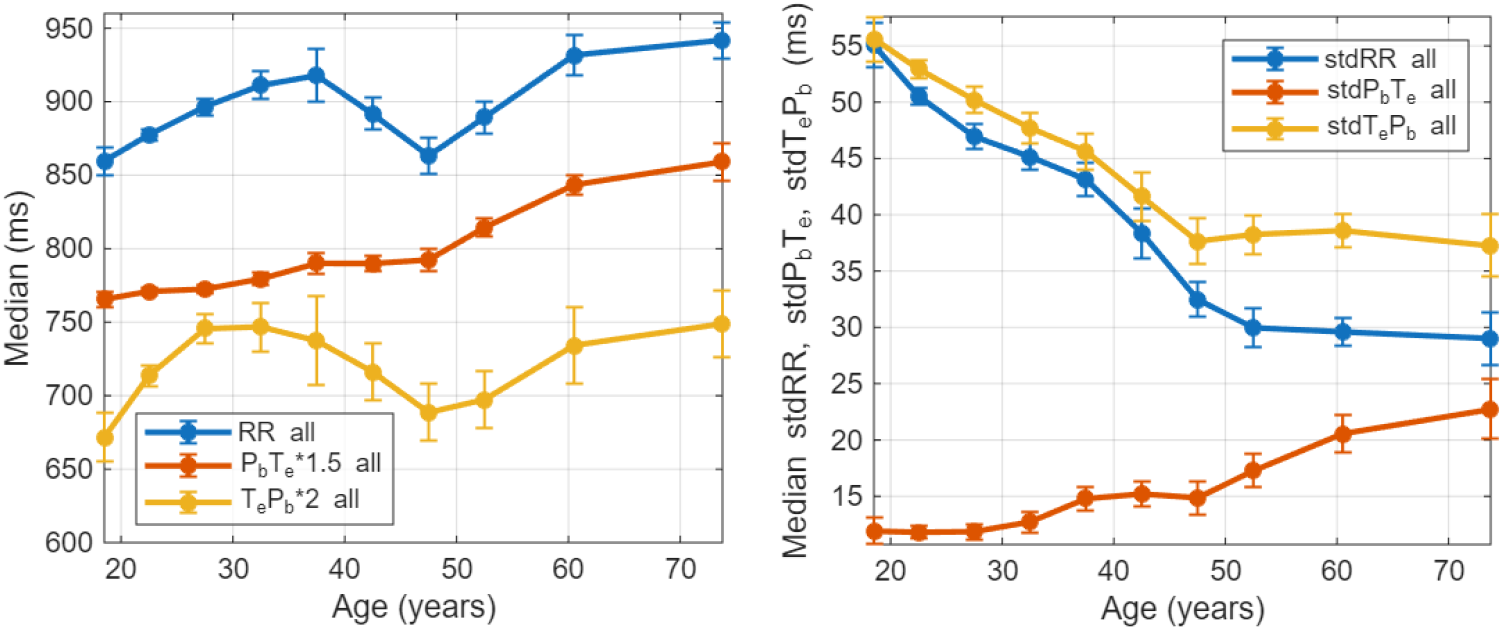
(a) Age variation of 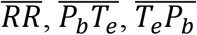 and their standard deviations. Scaling multiplication factors for 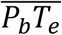 and 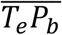 and their errors are 1.5 and 2.0, respectively. (b) Age variation of 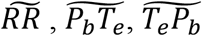. In all plots, the error bar length on each side indicates one standard deviation. All curves are smoothed with a 3-point triangular weighting function.

The active interval duration, 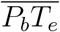, shows a progressive increase with age characterized by a larger slope after 50 years, as seen in Figure 1(a). The increase rate in median 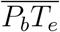 is 1.70 ms per year on average from 47.5 to 73.8 years, while the rate is substantially lower at 0.62 ms/yr from 18.5 to 47.5 years. Similar to 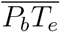, the 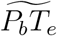 incremental rate of 0.3 ms/yr after 50 years old is three times that of the preceding age range. The isoelectric segment or idle interval, 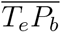, displays a similar age variation to 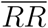. The minimum 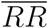, occurring at 47.5 yrs old, corresponds to the minimum in 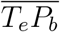. Cohort 1 has the largest 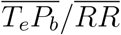 (43.3%) while Cohort 3 has the smallest at 38.1%. There is a strongly linear relationship between 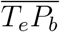 and 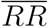, with the cross-correlation coefficient between the two parameters being about 0.95. 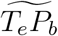, as seen in Figure 1(b), similarly decreases with age in tandem with 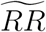, and is much larger than 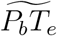, especially for younger subjects. Overall, it is evident that the age variations of 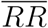 and 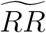 are primarily driven by 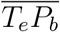 and 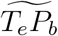, respectively. The characteristics of *RR, P*_*b*_*T*_*e*_, and *T*_*e*_*P*_*b*_ intervals indicate that 50 years is a pivotal transition age for the population represented in the Autonomic Aging dataset.

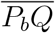, which represents the atrial conduction time, shows a gradual lengthening from young adulthood onward. This progressive lengthening is largely due to changes in 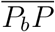, whereas 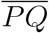 remains relatively stable over the entire age range from 20 to 70 years, as seen in Figure 2(a). Although the average rate of change in 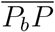 is merely 16 ms over a span of 55 years, it represents a nearly 50% relative change owing to its short baseline duration. 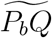 and 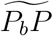, plotted in Figure 2(b), do not show significant changes before 50 years old but increase afterward.

**Figure 2.**
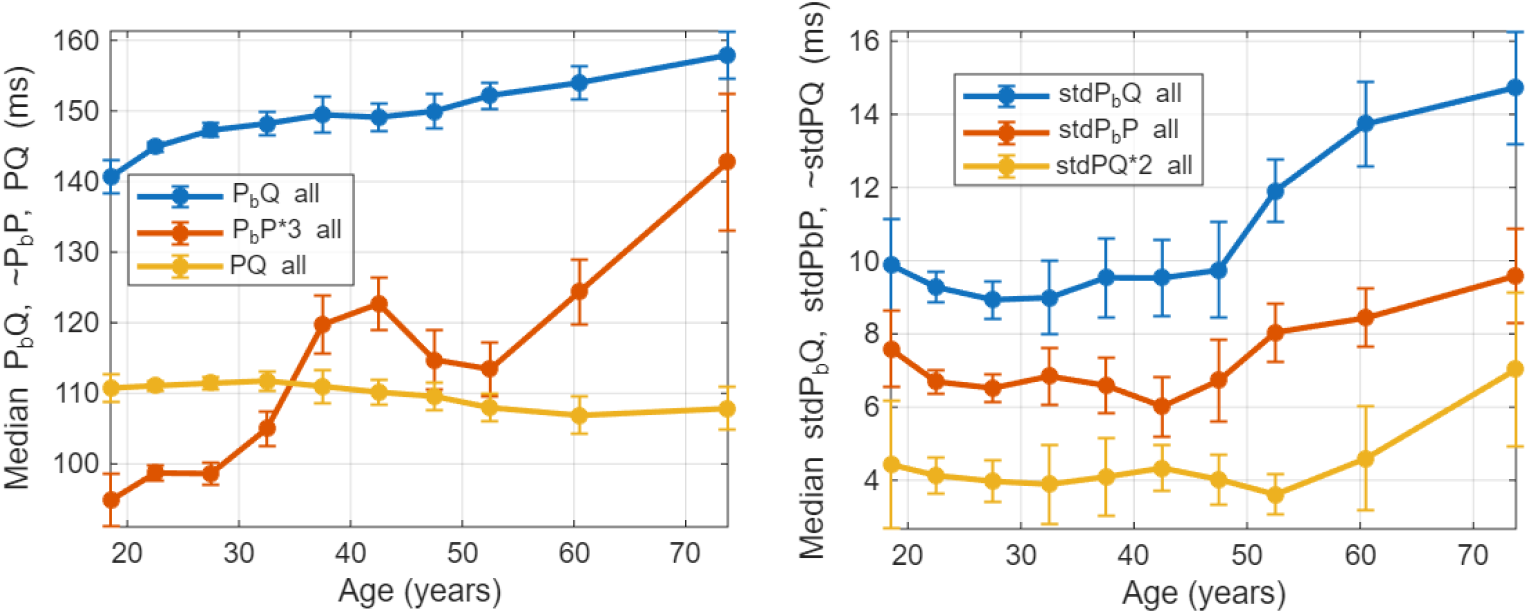
(a) Age variation of atrial intervals: 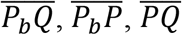 (b) Age variation of 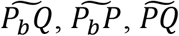. The scaling factors for 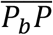 and 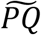 and their error bars are shown in the legend. All curves are smoothed with a 3-point triangular weighting function.

The ventricular interval durations and variabilities are plotted in Figure 3. 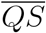, which is a good proxy for the ventricular depolarization time, shows a distinct aging effect: it steadily increases throughout the years with an accelerated rate after 50 yrs. The median 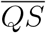 at 74 yrs is 50% longer than that at 20 yrs. In contrast, 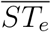, which is approximately the ventricular repolarization time, remains essentially constant with age. The age variation of the total ventricular conduction time, 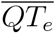, is largely caused by that of 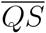. The variabilities of the three ventricle intervals exhibit faster changes after 50 yrs old than before. Overall, the impact of age on ventricular interval durations and their variabilities is similar to that on atrial intervals: the change over age in atrial and ventricular conduction times is primarily driven by the depolarization times, and the rate of change in interval lengths and variabilities accelerates after 50 years old.

**Figure 3.**
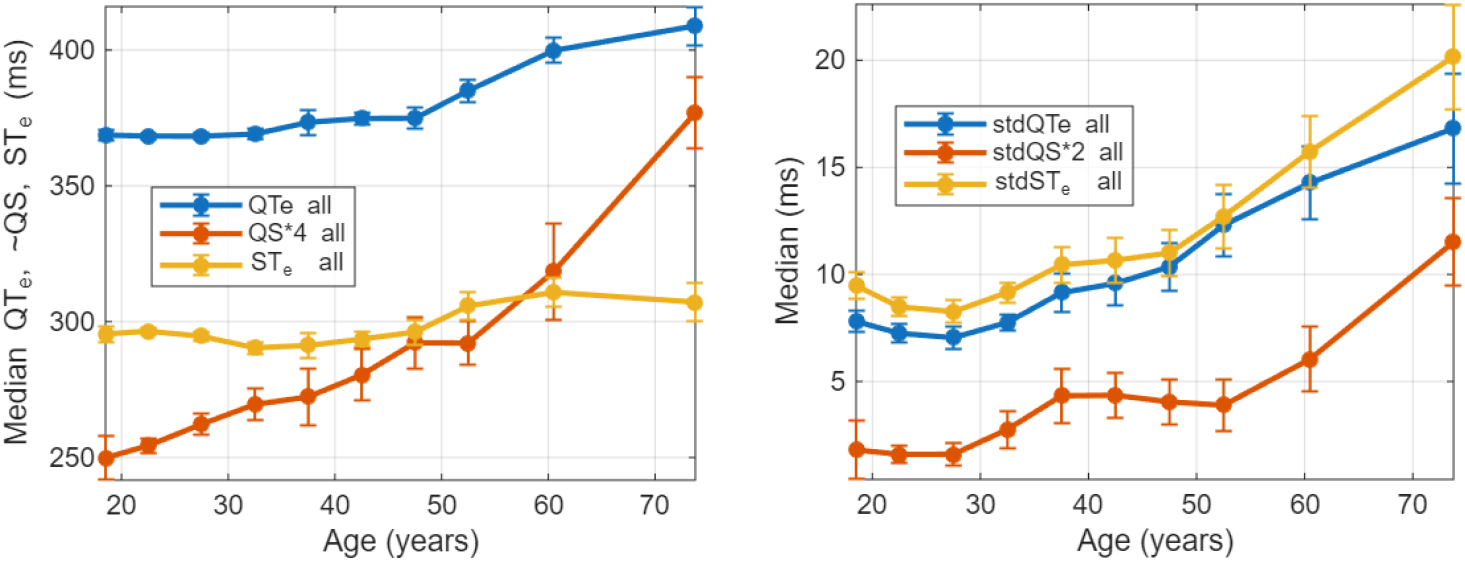
(a) Age variation of ventricle intervals: 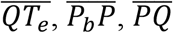. (b) Age variation of 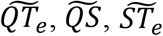. The scaling factors for 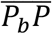, and 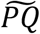 and their error bars are shown in the legend. All curves are smoothed with a 3-point triangular weighting function.

In summary, the analysis of interval lengths and their variabilities indicates that fifty is a critical transition age. Above this transition age, the active interval duration, 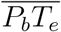, rises with age much faster than below. The change in 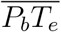 is primarily driven by modifications in the atrial and ventricular depolarization times. Variabilities in all atrial and ventricular intervals increase at a faster rate after 50 years old. 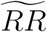 and 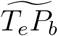 decrease at a steady rate from 20 to 50 years, but remain stable afterward. Previous studies also found that atrial and ventricular conduction times, as well as QRS duration, increase with age (Rabkin et al., 2016; Tan et al., 2016; Giovanardi et al., 2022; Ahmadi et al., 2023). Nevertheless, an important caveat is that the characteristics outlined in this section are derived from all subjects meeting the inclusion criteria. As discussed in the following, sex and BMI play crucial roles and must be factored in for a more accurate assessment of the characteristics.

Although the subjects in the Autonomic Aging dataset do not have known heart ailments, a larger deviation from the state of youth is a sign of aging and signifies a higher vulnerability to potential pathological changes. Aging does not appear to affect 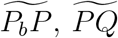, or 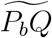 until 50 years of age when it starts to rise. The small variability of atrial intervals and the lack of change suggest that atrial arrhythmia should be relatively rare before 50 yrs, which is largely consistent with previous reports. For example, Lane et al. (2017) report that the incidence rate of atrial fibrillation in the age group of 55–64 yrs is nine times that observed in individuals younger than 54. Our analyses show that ventricular intervals and their variabilities increase more gradually with age, but with a faster rate around 55 yrs. These characteristics align with the studies by Sirichand et al. (2017), who report that ventricular ectopy and idiopathic ventricular arrhythmias increase gradually from mid-adulthood and rise sharply after about 60 yrs, and by Chow et al. (2012), who indicate that the most dramatic change occurs from about 60–70 years onward when left ventricular hypertrophy, diastolic dysfunction, and arrhythmias become common in the general population. It is generally understood that conduction speed is modified due to primary (idiopathic) fibrosis, congenital and secondary causes, with aging hastening the progressive loss of conduction fibers and calcification (Waller et al. 1993; Ali et al. 2024). Our results suggest that the change in atrial and ventricular conduction speed primarily occurs during the depolarization processes.

### 3.2. Effect of sex on ECG durations and their variabilities

In our study on the dependence of ECG parameters on sex and BMI, we contrast two age divisions: D1 for individuals from 18 to 29 yrs and D2 for those from 45 to 74 yrs, with the BMI threshold set at 25.5 kg/*m*^2^. The two age divisions and the BMI threshold were selected to balance the sample sizes (*n*) across the age, sex, and BMI partitions. Table 2 lists the mean age, mean BMI, and their standard deviations for eight partitions. We plot the cumulative distribution functions (CDF) of the nine interval durations for the four age and gender partitions with BMI ≤ 25.5 in Figure 4. In the legends, “b” is the BMI identifier, “M” and “F” are sex identifiers, while D1 and D2 denote the age divisions. The three numbers following “D1” or “D2” correspond to the median, mean, and standard deviation, respectively. Inter-sex p-values for D1 and D2 are labeled “p-v D1” and “p-v D2,” while “p-v M” and “p-v F” represent p-values between the two age divisions for each sex. These p-values are derived from two-sided tests for the null hypothesis that the means are equal.

**Table 2.**
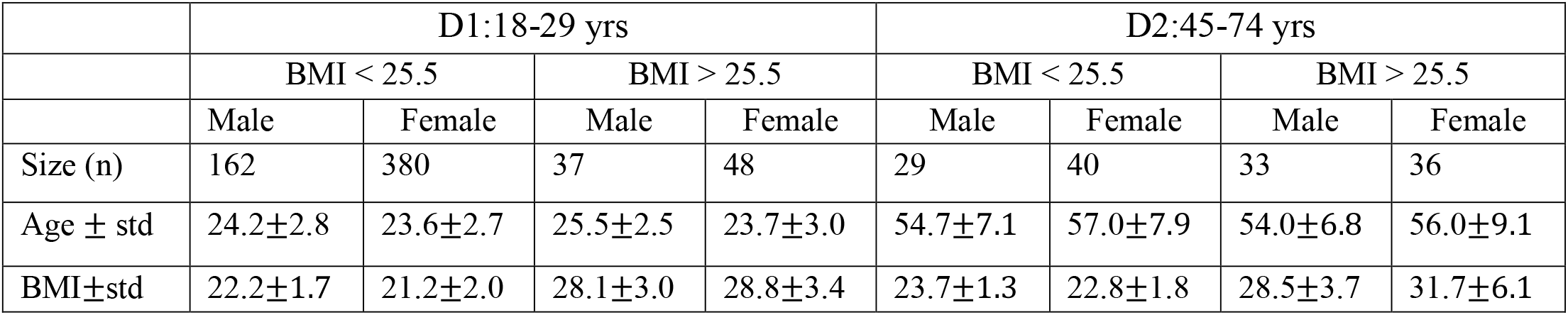
Basic statistics for gender and age partitions.

**Figure 4.**
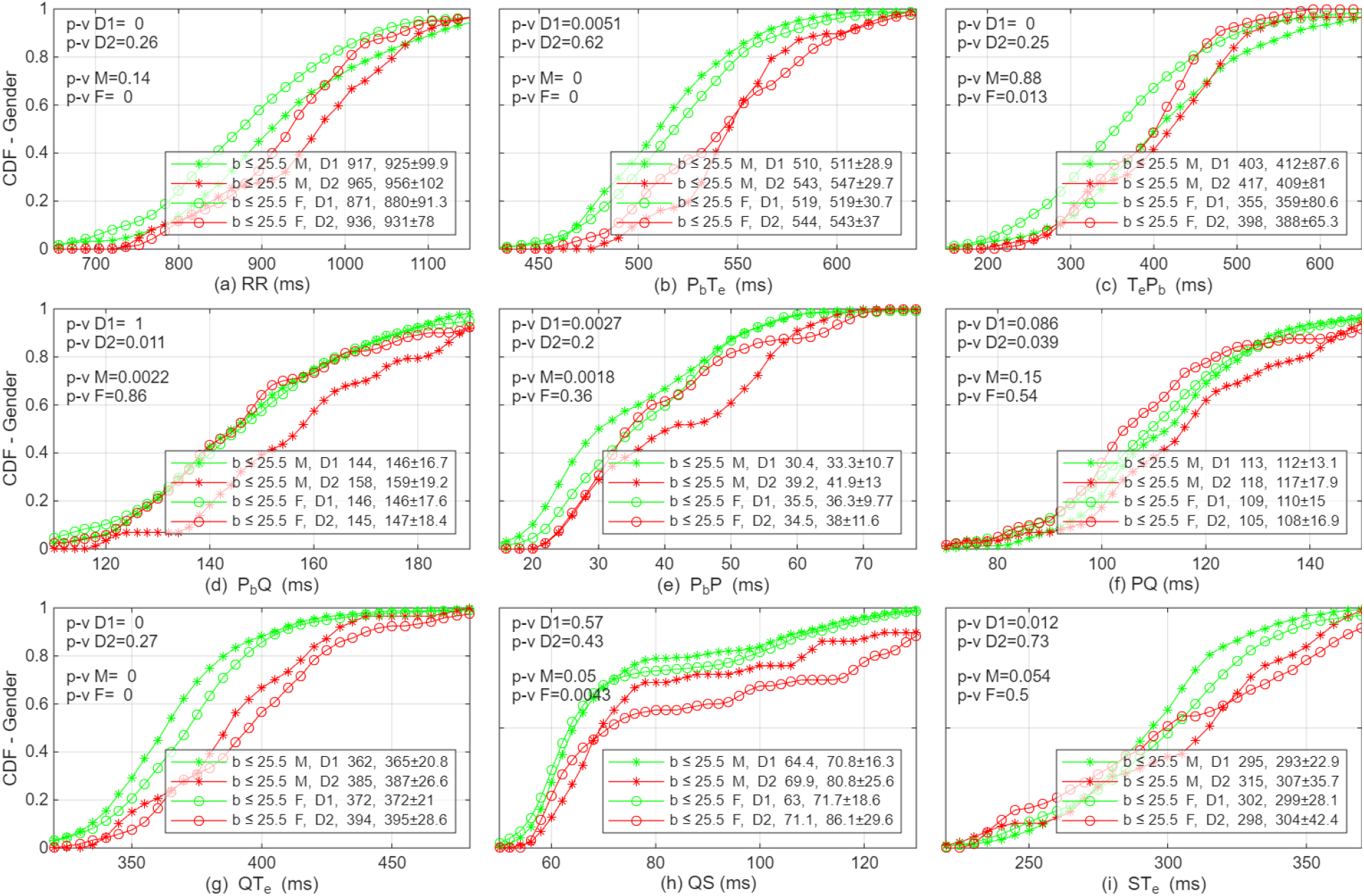
Cumulative distribution functions of 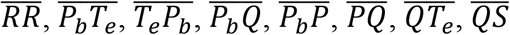, and 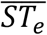 for sex and age partitions. “M” and “F” are for males and females, respectively. In the legend box, “D1” and “D2” are for 18-29 and 45-74 years of age. The three numbers following “D1” or “D2” are the median, mean, and standard deviation, respectively.

It is well known that female hearts beat faster due to their typically smaller size. This is confirmed in Figure 4(a), which shows that females’ 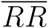 is shorter than males’ for individuals with low BMIs. For the younger age division, notable differences include: males’ median 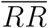 is 46 ms longer than females’, a difference entirely driven by men’s longer 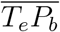 as their 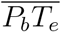 is shorter than women’s; there appears to be no significant sex difference in 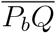 and 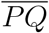, even though men tend to have a shorter 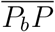 than women; men also exhibit a shorter 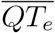, resulting from a shorter 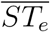. For the older age division, men’s median 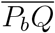 is 13 ms longer than women’s, primarily due to the formers’ longer 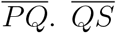 does not show an appreciable sex difference. The characteristics of age evolution from D1 to D2 (with an average age of from 24 to 56 years old) include: 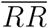 lengthens for both sexes, mainly due to increases in 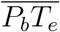; females’ 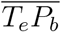 lengthens, while males’ remains relatively stable; all atrial intervals for men increase with age, whereas those for women do not change significantly; in the ventricles, all intervals for men increase, while for women, 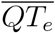 and 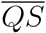 increase but 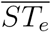 does not.

Previous studies in general show that men have a slower heart rate, a longer atrial conduction time, and a shorter ventricular conduction time (Macfarlane et al., 1994; Wu et al., 2003; Tan et al., 2016, Ngoc et al. 2025). While our study agrees with previous studies for the most part, differences exist. When the subjects are confined to low BMI, we do not see a significant difference between young women and men in atrial conduction time. While some previous studies (Macfarlane et al., 1994; Tan et al., 2016; Macfarlane, 2018) indicate that QRS is longer in men than women, we do not see a statistically significant sex difference in 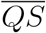.

Since individuals may possess unique heart interval patterns, the relative proportions of these subintervals may provide critical context for cardiac assessments. The top three rows of Figure 5 illustrate the ratios of the idle interval 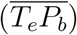, atrial conduction 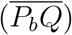, and ventricular conduction 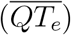 to the total cardiac cycle 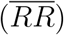, i.e., 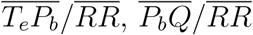, and 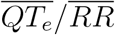, respectively. As discussed in the following section, 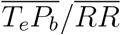 serves as an indicator of the heart’s capacity to function without requiring significant sympathetic nervous activity. In the younger age division, men exhibit a significantly larger 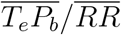 and smaller 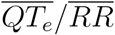 and 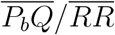 compared to young women, although these sex-based disparities become less pronounced in the older age division.

**Figure 5.**
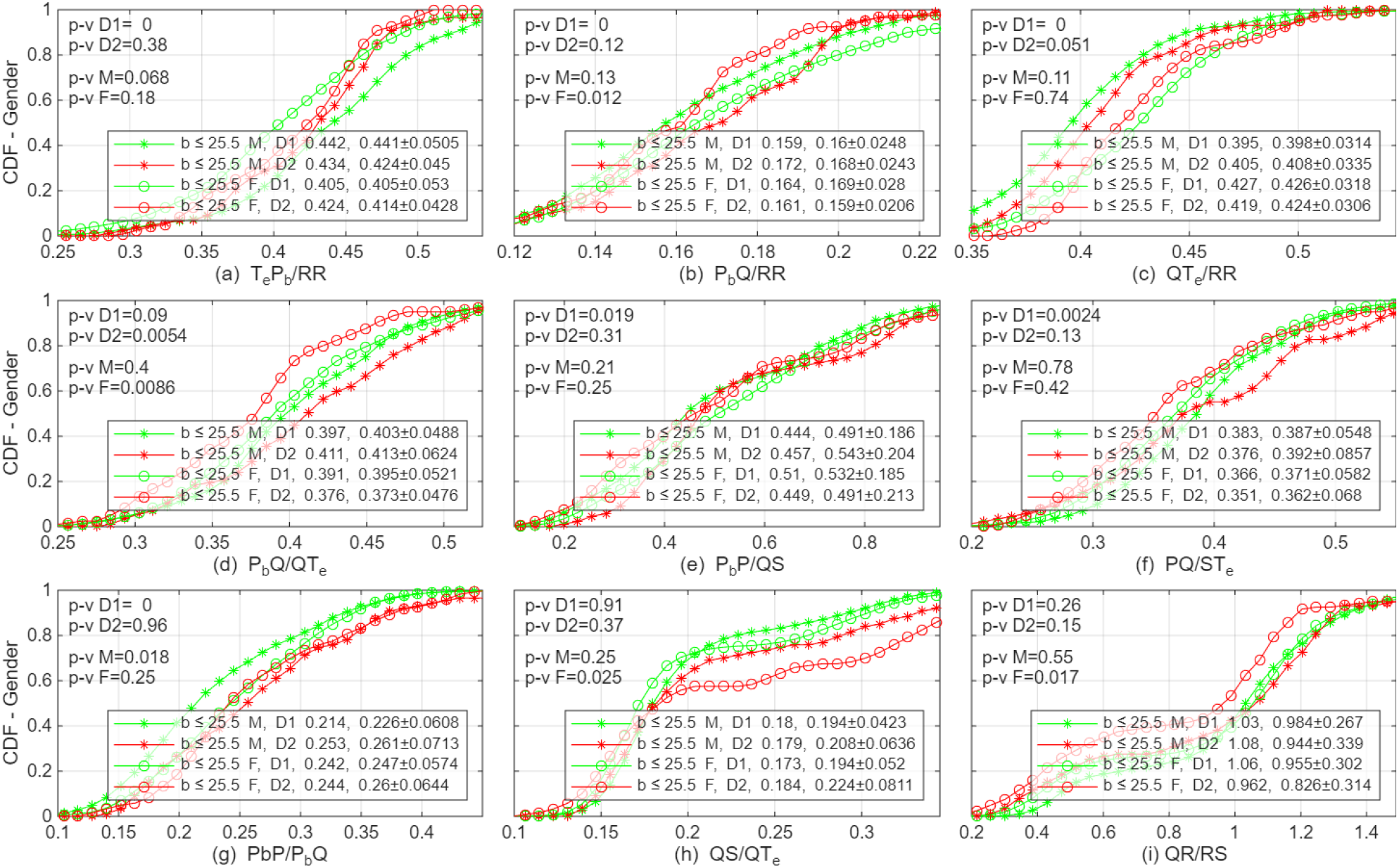
Cumulative distribution functions of 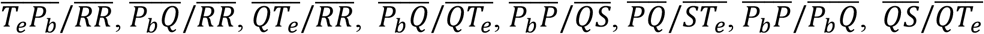, and 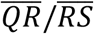 for sex and age partitions.

The second row of Figure 5 contrasts total conduction, depolarization, and repolarization times of the atria against those of the ventricles. These comparisons evaluate whether atrial and ventricular intervals change proportionally. Among younger subjects, no significant sex difference exists in the ratio of overall atrial to ventricular conduction time 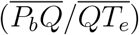; however, this ratio increases with age in men while decreasing in women. This divergent age-related evolution results in a significant sex difference within the older population. Despite the negligible sex difference in 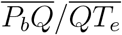 for younger individuals, there are marked differences in depolarization and repolarization ratios: younger men display a smaller 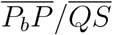 and a larger 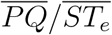 compared to younger women.

The ratios 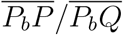 and 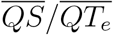 quantify atrial and ventricular depolarization times relative to their respective conduction durations. Younger men possess a smaller 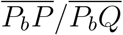 than younger women, but this difference diminishes with age. Specifically, the 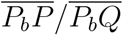 ratio in men tends to increase over time, whereas it remains largely stable in women across the two age divisions. No significant sex difference is observed for 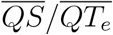 in either age division. However, 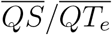 increases as women age, while men do not experience a similar shift. Finally, the 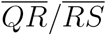 ratio, shown in Figure 5(i), characterizes the symmetry of the QRS complex. The median 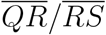 is approximately 1, with values for older women slightly lower.

The variabilities corresponding to the nine intervals plotted in Figure 4 are shown in Figure 6. In the younger age division, men’s 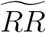 is slightly larger than women’s, while the variabilities of all other intervals do not exhibit significant gender differences. In the older age division, women’s 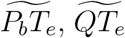, and 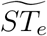 are larger than men’s with a p-value less than 5%. 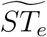 may potentially drive the variabilites of *QT*_*e*_ and *P*_*b*_*T*_*e*_, which contain the *ST*_*e*_ interval. Numerous studies have investigated gender differences in 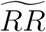. A meta-analysis by Koenig and Thayer (2016) indicates that females generally possess an overall significantly lower 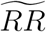, notwithstanding certain studies that do not corroborate this characteristic (e.g., Murata et al., 1992). Although the Autonomic Aging dataset shows our result indicates that younger women have a lower 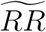 than younger men, the primary factor influencing 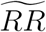 is age.

**Figure 6.**
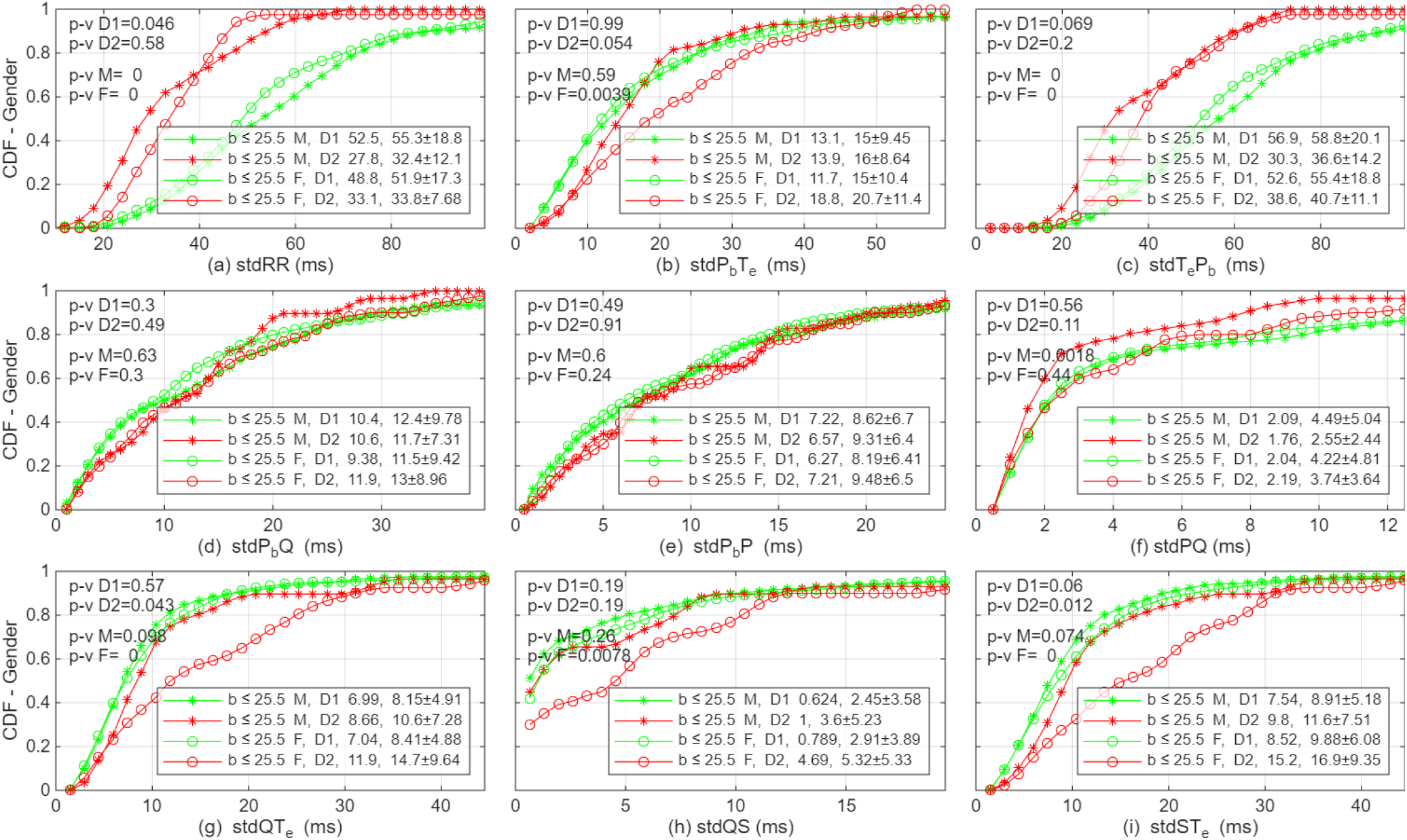
Cumulative distribution functions of 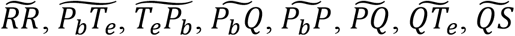, and 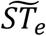 for sex and age partitions.

If we assume that younger hearts are intrinsically healthier, the increase in atrial interval durations in men as they age may suggest they are more prone to developing atria-related health issues than women. This aligns with the age-adjusted atrial fibrillation (AF) rate, which is 1.5–2 times higher in men than in women (Ko et al., 2016). While 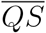 in both sexes increases with age, men’s 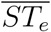 increases as well while women’s does not show a change. Older women, conversely, exhibit larger variabilities in the ventricular intervals. This gender disparity may potentially be associated with the distinct ventricular diseases that the two sexes typically experience.

### 3.3. Effect of BMI on heart intervals and their variabilities

We divide the BMI into two groups (≤ 25.5 kg/*m*^2^ and *>* 25.5 kg/*m*^2^) using the same age divisions as previously described. For this analysis, the two sexes are pooled due to small sample sizes in certain partitions. The results are weighted toward females owing to their larger representation. The interval durations for age- and BMI-based partitions are plotted in Figure 7, similar to Figure 4 for the sex partition. As illustrated in Figure 7(a), 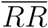 is shorter for individuals with high BMI, especially in the older group. While the heart rate of low-BMI individuals decreases with age, that of high-BMI individuals increases, imposing a significant burden. For one unit of BMI increase, the average heart rate increases by 0.97 bpm for the older division, while it is 0.19 bpm for the younger division. As seen in the above, the 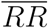 characteristics are mirrored in 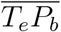. BMI does not affect 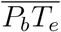 in either age group. BMI does not significantly influence the six atrial and ventricular intervals in the older division, although high BMI corresponds to somewhat longer 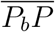 and 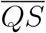. In the younger division, high BMI is associated with shorter 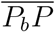 and 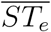, but longer 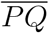 and 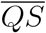. Because the atria and ventricular depolarization and repolarization times shift in opposite directions, the total conduction times remain largely insensitive to BMI.

**Figure 7.**
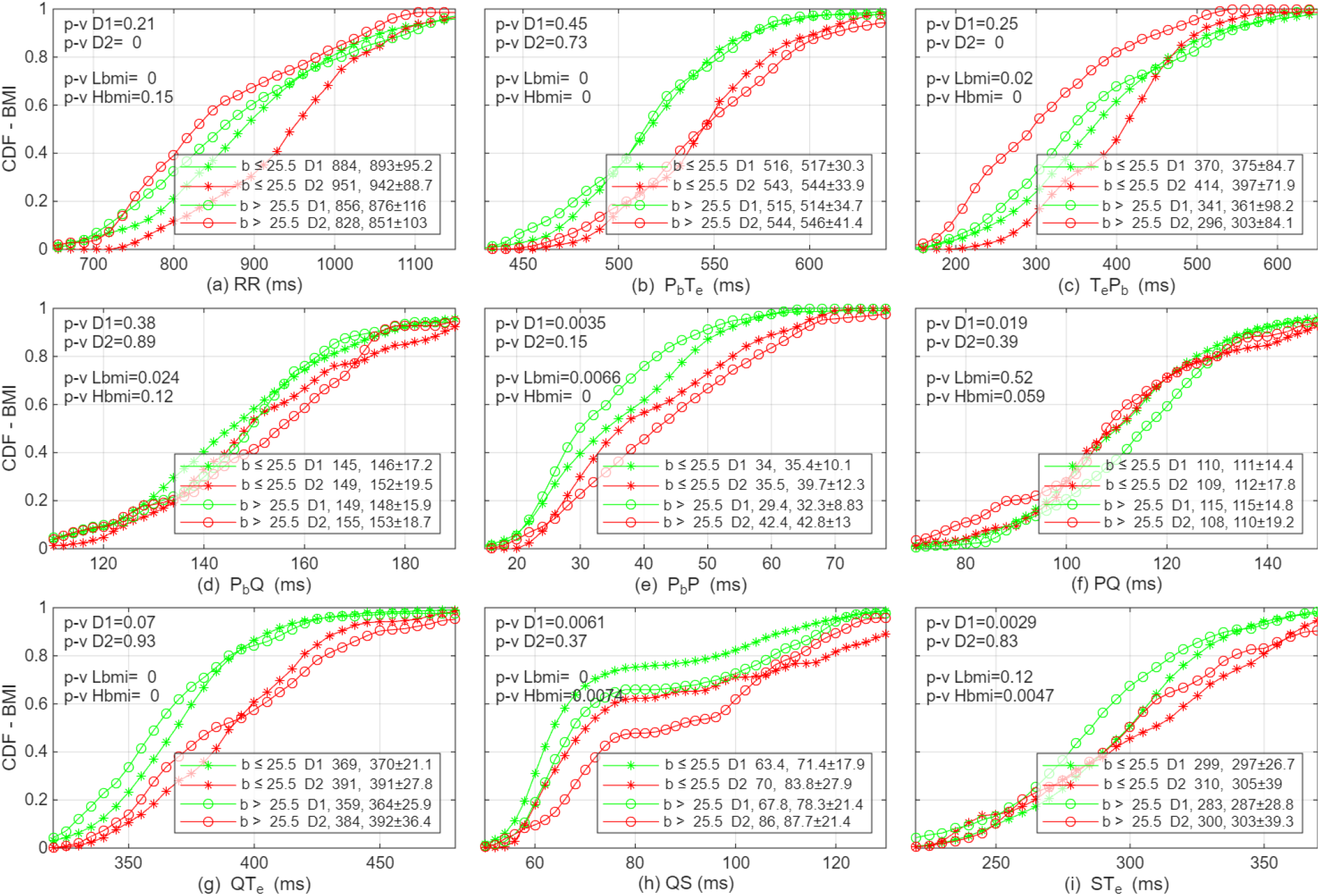
Cumulative distribution functions of 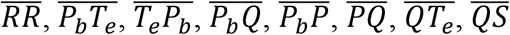, and 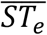 for four age and BMI partitions.

The relative intervals based on BMI partitions are shown in Figure 8. The BMI effect on 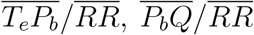, and 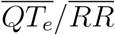 is most pronounced in the older age division. Older people with higher BMI have the smallest 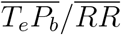, but the largest 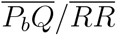 and 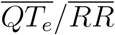. The differences in these ratios for the younger age division are less significant. The hearts of older and higher-BMI individuals are in the idle state 35% of the time, compared to 42% for their low-BMI counterparts. The durations of atrial and ventricular conduction times relative to one’s own 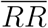 do not change with age for low-BMI people, while they increase significantly for the high-BMI population. For the older division, an increased BMI does not change the ratios between the durations of the atrial and ventricular intervals. It appears to increase 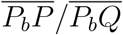 and 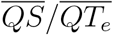 but decrease 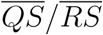, even though the statistical significance is weak due to small sample sizes. For the younger age division, high BMI decreases 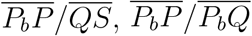, and 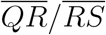, but increases 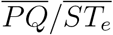 and 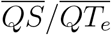.

**Figure 8.**
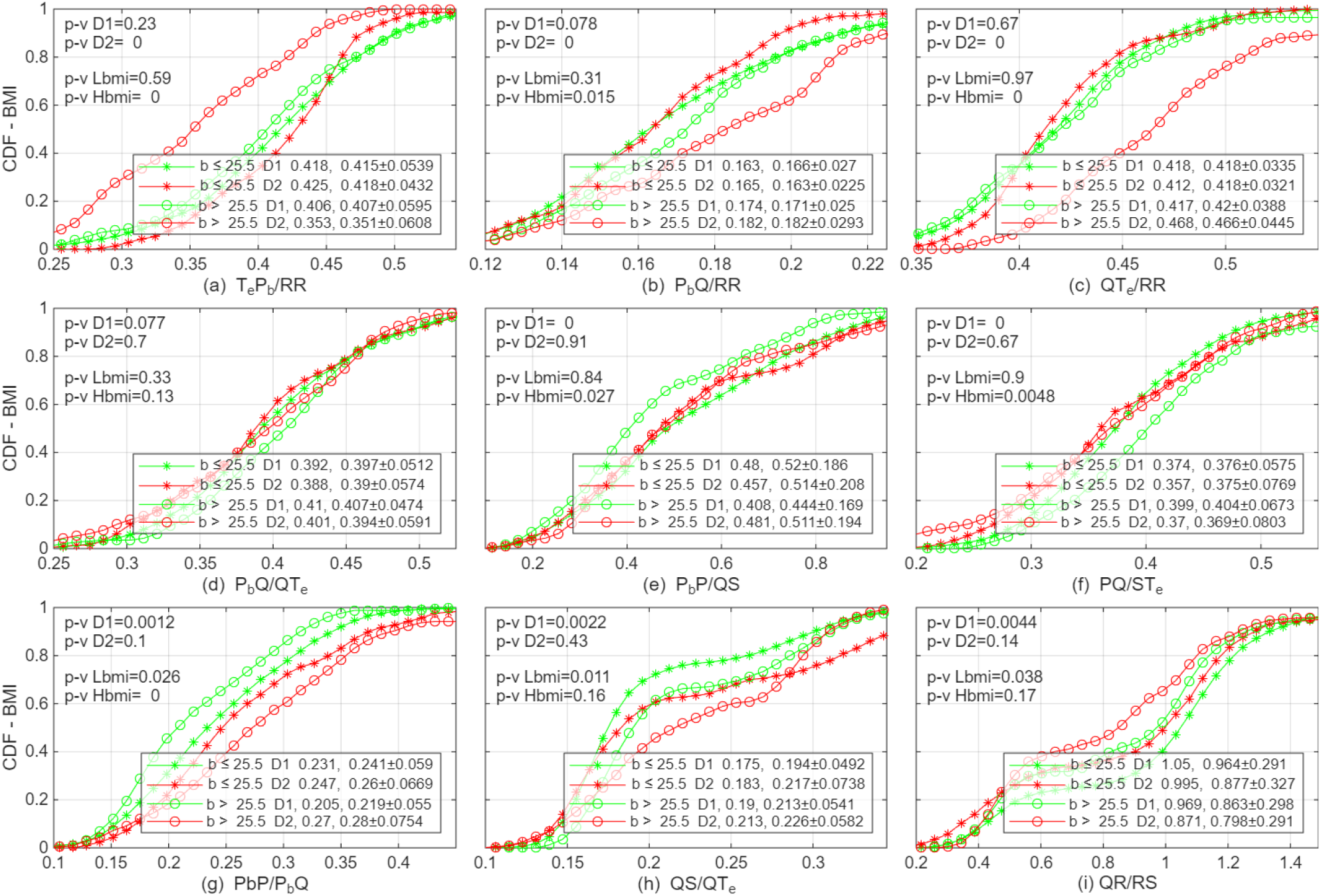
Cumulative distribution functions of 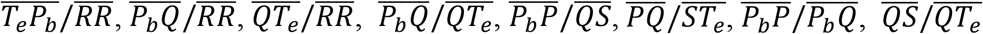, and 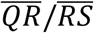 for BMI and age partitions.

The variability plots based on BMI and age partitions are shown in Figure 9. BMI does not have a significant impact on the variabilities of all nine intervals for the younger age division. Of statistical significance are 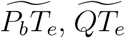, and 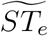 in the older age division. For those three parameters, higher BMI is associated with greater variability. As seen previously, 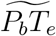, 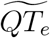, and 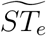 are larger in low-BMI older women than in older men. The BMI differences in the older population for these three parameters are mostly contributed by men. The variability of *ST*_*e*_ 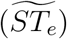 is of particular importance as it is a component of both *P*_*b*_*T*_*e*_ and *QT*_*e*_.

**Figure 9.**
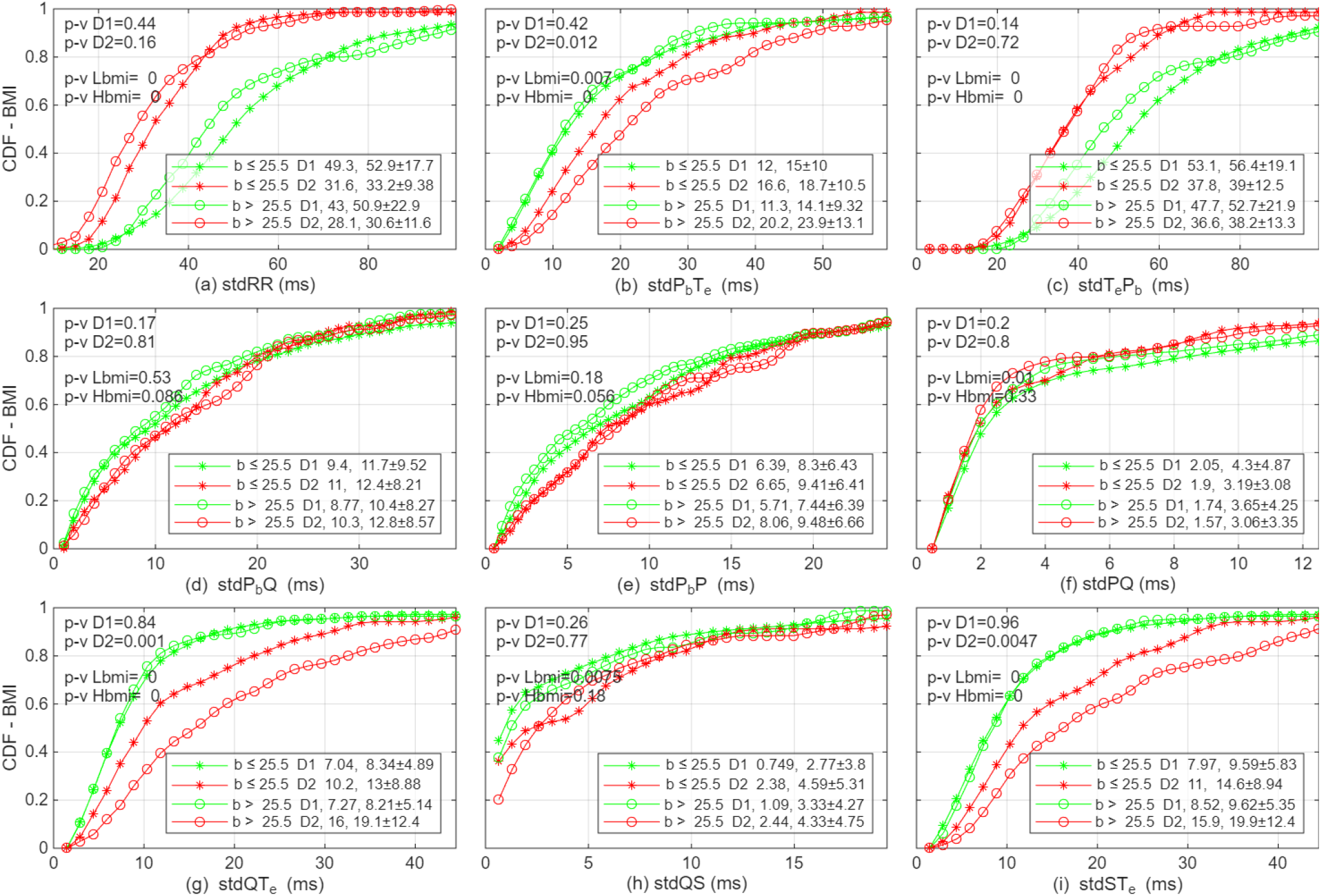
Cumulative distribution functions of 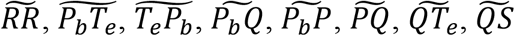, and 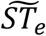 for BMI and age partitions.

Our results largely agree with previous studies demonstrating that a higher BMI increases the heart rate and the atrial conduction time (Ardissino et al., 2023; Yang et al., 2024). While some previous studies indicate that a higher BMI also increases the ventricular conduction time, there are also reports that do not show such an association. Our analysis shows that the ventricular conduction time is largely controlled by age rather than by BMI. While previous reports are inconclusive regarding the QRS duration, our study indicates that higher BMI is correlated with increase in QS duration.

It should be noted that the BMI statistics presented here are limited by the small sample sizes in certain partitions. In general, an observation of a significant difference is more conclusive than an observation that does not show a significant difference. Taking Figure 9(b) as an example, it is a relatively firm result that BMI exerts a statistically significant difference on 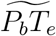 for older people. The statistically less significant difference for the younger age division can be due to the smaller sample sizes or insufficient spread in our low and high BMI definitions.

## 4. DISCUSSION

While the heart is a complex organ, how it modulates cardiac output is analogous to the conceptually simple pulse-width modulation (PWM) power system, which is widely used in engineering, such as in a radar, to adjust output power. The output power in a PWM system is modulated by the duty cycle, i.e., the ratio of the “on” time to the pulse period. In the heart, the “on” time is when the electrical signal propagates through the atria and ventricles 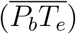, i.e., the active interval. The “off” time or idle interval corresponds to the isoelectric segment after ventricular repolarization and before the next atrial depolarization 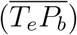, which is commonly referred to as the *TP* segment. The resting heart functions as a PWM system due to its small 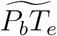 and large 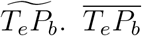 is often taken as part of the ventricular repolarization process. We emphasize the importance of the idle interval in its own right and its functional specificity. Presence of idle time allows the heart to modulate its output with minimal adjustment to the active conduction period in the parasympathetic-dominant mode. A heart with a longer 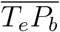 or 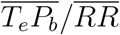 is in a more relaxed state, possessing greater “reserved power” before transitioning to sympathetic-dominant mode where 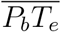 must be shortened to meet the body’s increasing demand.

The most notable differences between sexes and BMI groups for the resting hearts are manifested in the idle interval. Young men typically have a significant advantage in idle time over young women. As shown in Figures 4 and 5, the average 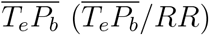 is 403 ms (43.5%) for young males compared to 360 ms (40.5%) for young females. High BMI decreases both 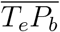 and 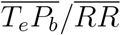, an effect particularly pronounced in the older population as seen from Figures 7 and 8. Stratification of age, BMI, and sex for 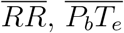, and 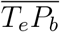 is illustrated in Figure 10 for seven age bins. 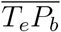 is shorter in the higher-BMI groups for both sexes. The difference between the two male groups in 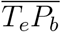 and 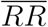 is far more substantial than that between the two female groups. The intra-sex difference in 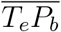 for males exhibits a strong age dependence. In contrast to the divergence in 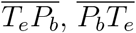 shows small differences among the four partitions. 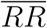 exhibits features similar to 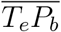 but trends upward for low BMI groups owing to 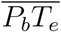 increasing with age.

**Figure 10.**
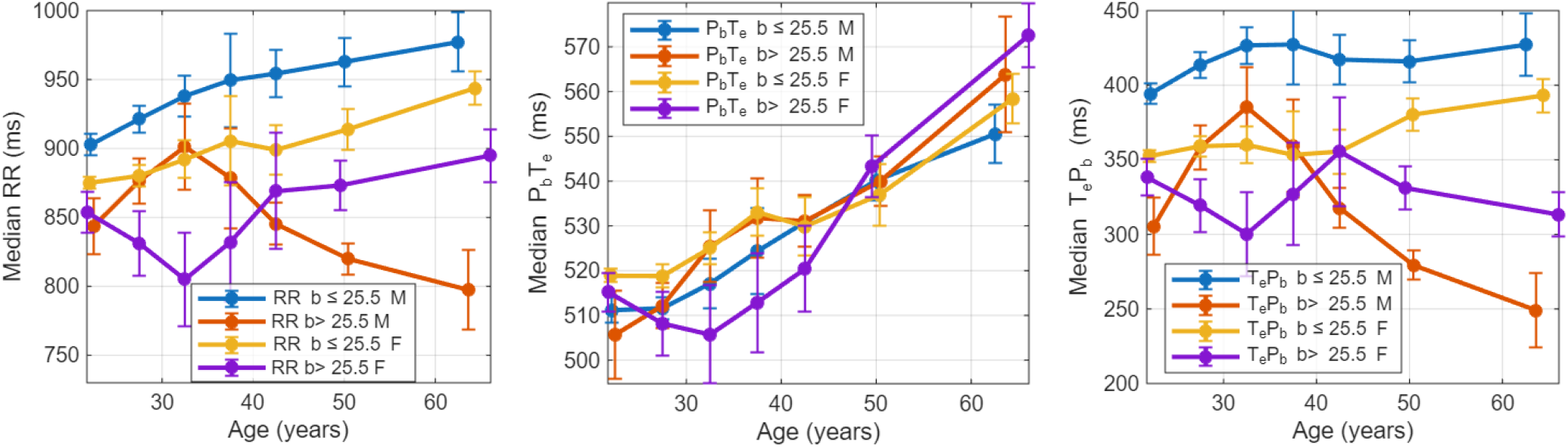
Variations of 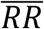 (left), 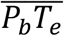 (middle), 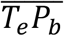 (right) with age for BMI and sex partitions.

The distinct age evolutions of 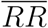 for the four partitions shown in Figure 10(a) illustrate the importance of stratification. The higher average heart rate near 50 yrs of age seen in Figure 1 is associated with a higher average BMI. For low BMI people, 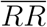, similar to 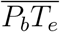, increases monotonically with age. The male-female 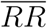 difference for the low BMI groups is about 50 ms, which translates to about 3.5 beats per minute faster for women, consistent with the result reported by Quer et al. (2020). A striking feature of Figure 10 is that high-BMI males’ age variation of 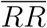, driven by 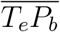, drops sharply after 35 years of age, whereas it increases for the other three partitions. The 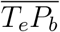 and 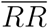 characteristics indicate that being overweight stresses the male hearts more than it does the female hearts, especially after 35 years old.

It is generally held that a greater resting heart rate variability is a sign of youth and better health. What causes the greater variability, and to what extent it acts as a health indicator need further clarification and study. The variabilities of the active interval 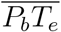 and its subintervals, such as 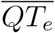, tend to increase as people age. Older people’s more frequent firing of sympathetic nerves increases the variability of the active interval and subintervals. An important characteristic of a younger and healthier heart is its steady timing of the active interval and subintervals. The heart rate variability is primarily driven by the variability of the idle time 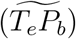. A general explanation for younger people’s higher 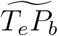 and 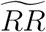 is that their autonomic nervous system is more responsive. Another plausible reason that deserves further study is that younger people simply have a larger variation in their instantaneous physiological need for cardiac output. As the modulation of 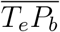 and 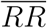 is most directly associated with the need for cardiac output, the durations and variabilities of *T*_*e*_*P*_*b*_ and *RR* are not as direct heart health indicators as those of *P*_*b*_*T*_*e*_ are. Figure 11 shows the age variations of 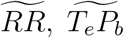, and 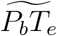 for the four sex-BMI partitions. If 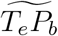 or 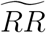 were a universal indicator of a younger or healthier heart, males with a higher BMI would have healthier hearts than those with a lower BMI at older ages, and people in their 70s would have an equally healthy heart as in their 50s. The usefulness of 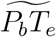 as an age and health indicator is indicated by the fact that older and higher-BMI individuals consistently exhibit higher 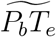 values. It is striking to see that high-BMI males in their thirties have elevated 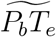 values exceeding those of low-BMI males in their sixties. The gap between high-BMI and low-BMI females is far smaller than that in males. In 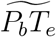, we again see that BMI impacts males far more than it does females.

**Figure 11.**
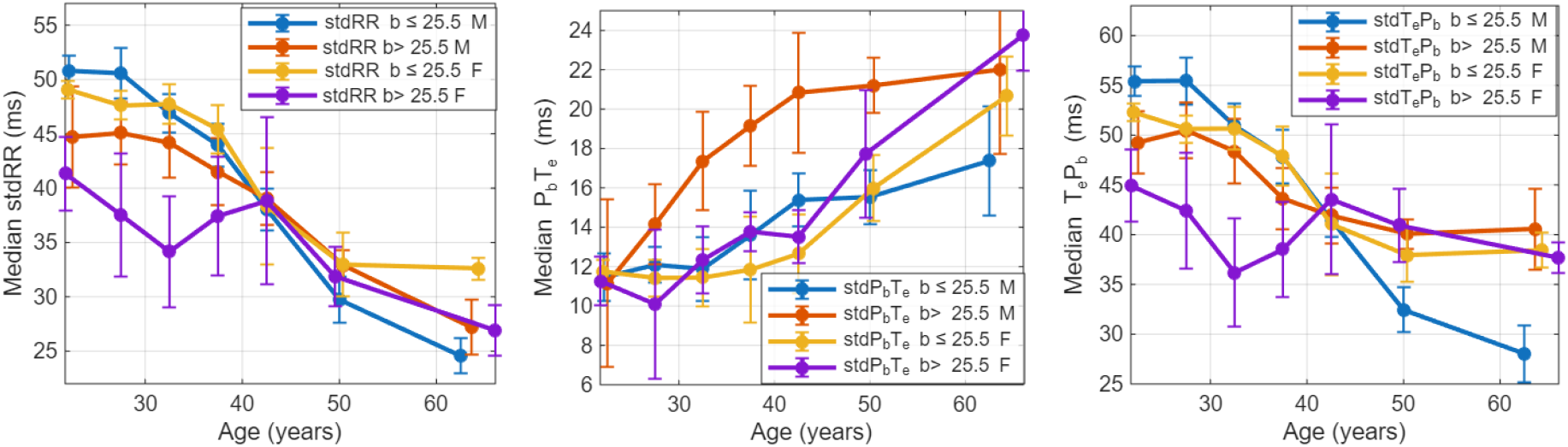
Variations of 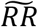 (left), 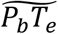 (middle), 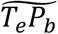(right) with age for BMI and sex partitions.

The elevated 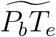 of high BMI males suggests that they are more susceptible to arrhythmia than other groups. Figure 12 further shows the atria and ventricular conduction times and their variabilities. High-BMI males have larger atrial and ventricular variabilities (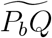 and 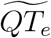) than low-BMI males. Most noticeably, their 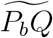, like their 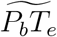, started to be highly elevated around 30 years old. A higher BMI, however, does not appear to affect females’ 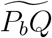 throughout the years but increases 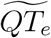 around 40-50 years old. For males, BMI emerges as a more dominant factor than age in the elevation of atrial conduction variability 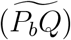. These features are consistent with previous reports (Singleton et al., 2020; Peng et al., 2025; Tan et al., 2025) that high BMI males have a much higher AF incidence rate.

**Figure 12.**
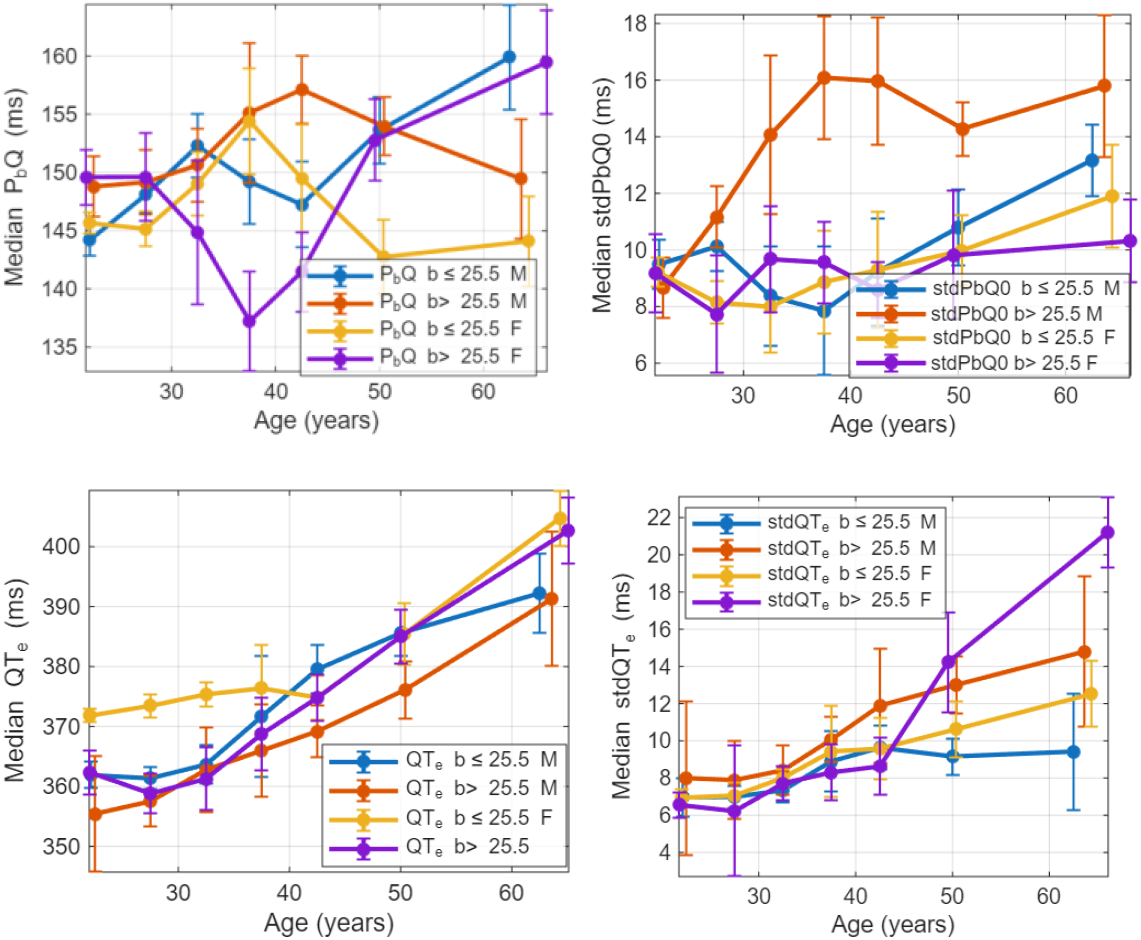
Variations of 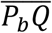 (top left), 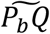 (top right), 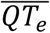 (bottom left) and 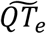 (bottom right) with age for BMI and sex partitions.

## 5. LIMITATIONS

The study would benefit from a larger sample size to be more statistically robust. The number of subjects is not even in age, sex, and BMI partitions.

## 6. SUMMARY AND CONCLUSION

The most salient features of our analyses are summarized in Figure 13. A cardiac cycle can be classified hierarchically into an active interval, comprising atrial and ventricular conduction times, and an idle interval, which modulates heart output. The characteristics of the *RR* interval are highly correlated with those of the idle interval, *T*_*e*_*P*_*b*_, for a resting heart. A healthy heart endeavors to maintain the stability of the active interval, *P*_*b*_*T*_*e*_, and its subintervals. Such a configuration allows the modulation of cardiac output without the need to adjust the operation of the atria and ventricles. This paradigm parallels that often used in engineering systems for simplicity, reliability, and durability. In healthy individuals at rest, the most pronounced sex and BMI differences are observed in the duration and variability of *P*_*b*_*T*_*e*_.

**Figure 13.**
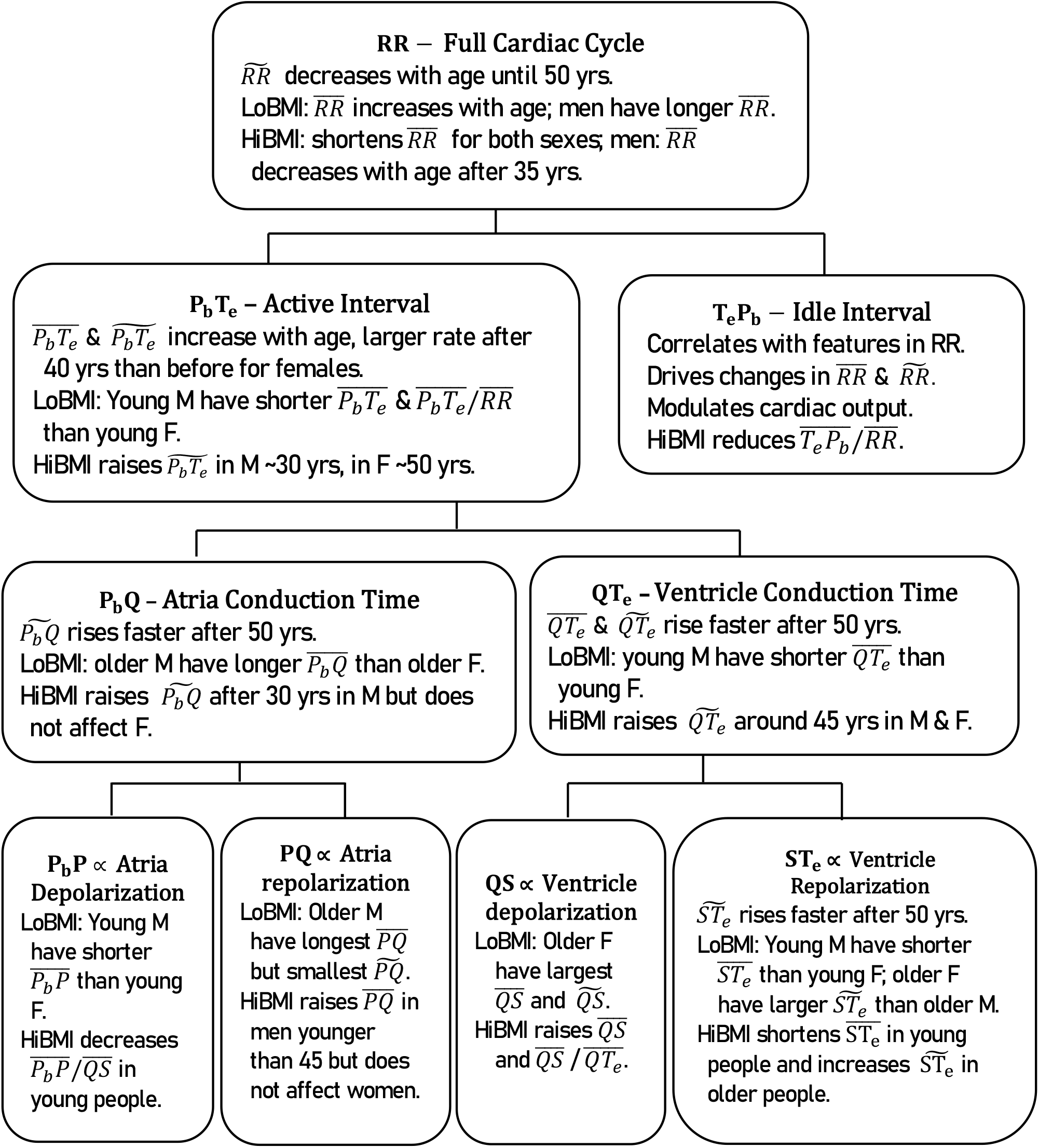
Major age, BMI and sex characteristics of ECG intervals. LoBMI and HiBMI are for low-BMI and high-BMI; M and F are for males and females, respectively.

Our results suggest that the heart undergoes its most pronounced changes around age 50 when the population is considered as a whole. 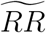 and 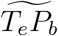 decrease with age until 50 yrs when they remain largely constant afterward. A decrease in 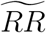 and 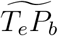 may not directly signal the degeneration of the heart as often thought. The degenerative signs lie in changes in the durations and variabilities of the active and its component intervals, particularly those associated with depolarizations, as summarized in Figure 13.

We have analyzed the sex differences for low-BMI individuals across two age divisions: 18–29 years and 45–74 years. For the younger age division: the cardiac period 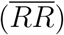 **is** longer and the active interval duration 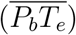 is shorter in males; the longer 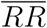 in males stems from their longer 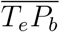, while their shorter 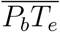 is mostly due to shorter 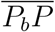 and 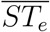; no significant sex difference is observed in the variabilities for all nine intervals examined. For the older age division, the sex differences for 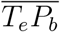 and 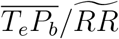 are smaller than those observed in the younger division. Older males have a significantly longer atrial conduction time, whereas older females exhibit a larger variability in ventricular conduction time.

The effect of BMI was examined using binary classification (≤ 25.5 kg/*m*^2^ for low BMI and *>* 25.5 kg/*m*^2^ for high BMI), with the same two age divisions as in the sex analysis. For the younger division: high-BMI subjects have longer 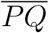 and 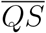, but shorter 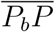 and 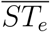; BMI does not significantly affect the variability of any of the nine intervals examined. For the older division: high BMI shortens the idle time, which is responsible for their accelerated heart rate; high BMI may also result in longer 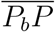 and 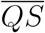, though the statistical significance is not strong due to small sample sizes; high BMI is correlated with greater variability in ventricular repolarization time, which may cause the increased variabilities in ventricular conduction time and the active interval.

Of consequence from the age, BMI, and sex stratification analysis is that high BMI has a much greater impact on males than on females. The heart rate of high-BMI males starts to accelerate in the early 30s, whereas it continues to decelerate in other BMI/sex partitions. In the early 60s, the median heart rate of males with an average BMI of 28.3 kg/*m*^2^ is 23% faster than the 61.4 bpm for males with an average BMI of 23.9 kg/*m*^2^. High BMI in males leads to greater variability in atrial conduction time and active time around 30 years of age, and in ventricular conduction time around 40 years of age. In females, high BMI also increases heart rate at all ages, but not as dramatically as in males after age 40. High BMI in females increases the variability of ventricular conduction time, but not that of the atrial conduction time. The varying effects of BMI across the two sexes warrant further studies with larger sample sizes and across different races.

## Data Availability

All data produced in the present study are available upon reasonable request to the authors

## Acknowledgement

The author would like to thank A. Schumann, K. Bär for making the Autonomic Aging dataset available for public access.

## Data Availability

The ECG recordings used in this study are available at https://physionet.org/content/autonomic-aging-cardiovascular/1.0.0/.

## REFERENCES

1. Abi-Gerges, N., Philp, K., Pollard, C., Wakefield, I., Hammond, T., & Valentin, J. (2004). Sex differences in ventricular repolarization: from cardiac electrophysiology to Torsades de Pointes. Fundamental & Clinical Pharmacology, 18., 10.1111/j.1472-8206.2004.00230.x.

2. Ahmadi, P., Afzalian, A., Jalali, A., Sadeghian, S., Masoudkabir, F., Oraii, A., Ayati, A., Nayebirad, S., Pezeshki, P., Tokaldani, M., Shafiee, A., Mohammadi, M., Sanei, E., Tajdini, M., & Hosseini, K. (2023). Age and gender differences of basic electrocardiographic values and abnormalities in the general adult population; Tehran Cohort Study. BMC Cardiovascular Disorders, 23. 10.1186/s12872-023-03339-z.

3. Ali, Z. S., Bhuiyan, A., Vyas, P., Miranda-Arboleda, A., Tse, G., Bazoukis, G., Burak, C., Abuzeid, W., Lee, S., Gupta, S., Eshfahani, M. A. M., & Baranchuk, A. (2024). PR prolongation as a predictor of atrial fibrillation onset: A state-of-the-art review. Current Problems in Cardiology, 10, issue 4. 10.1016/j.cpcardiol.2024.102469.

4. Ardissino, M., Patel, K., Rayes, B., Reddy, R., Mellor, G., & Ng, F. (2023). Multiple anthropometric measures and proarrhythmic 12-lead ECG indices: A mendelian randomization study. PLOS Medicine, 20. 10.1371/journal.pmed.1004275.

5. Carbone, V., Guarnaccia, F., Carbone, G., Zito, G., Oliviero, U., Soreca, S., & Carbone, F. (2020). Gender differences in the 12-lead electrocardiogram: clinical implications and prospects, Ital J Gender-Specific Med 2020;6(3):126–141. 10.1723/3432.34217.

6. Cavarretta, E., Sciarra, L., Biondi-Zoccai, G., Maffessanti, F., Nigro, A., Sperandii, F., Guerra, E., Quaranta, F., Fossati, C., Peruzzi, M., Pingitore, A., Stasinopoulos, D., Rigby, R., Adorisio, R., Saglietto, A., Caló, L., Frati, G., & Pigozzi, F. (2022). Age-Related Electrocardiographic Characteristics of Male Junior Soccer Athletes. Frontiers in Cardiovascular Medicine, 8. 10.3389/fcvm.2021.784170.

7. Chow, G., Marine, J., & Fleg, J. (2012). Epidemiology of arrhythmias and conduction disorders in older adults. Clinics in geriatric medicine, 28, 4, 539–553. 10.1016/j.cger.2012.07.003.

8. Dykiert, I., Kraik, K., Jurczenko, L., Gać, P., Poreba, R., & Poreba, M. (2024). The Effect of Obesity on Repolarization and Other ECG Parameters. Journal of Clinical Medicine, 13. 10.3390/jcm13123587.

9. Fisher, J., Young, C., & Fadel, P. (2015). Autonomic adjustments to exercise in humans. Comprehensive Physiology, 5 2, 475–512. 10.1002/cphy.c140022.

10. Giovanardi, P., Vernia, C., Tincani, E., Giberti, C., Silipo, F., & Fabbo, A. (2022). Combined Effects of Age and Comorbidities on Electrocardiographic Parameters in a Large Non-Selected Population. Journal of Clinical Medicine, 11. 10.3390/jcm11133737.

11. Goldberger, A., Amaral, L., Glass, L., Hausdorff, J., Ivanov, P. C., Mark, R., … & Stanley, H. E. (2000). PhysioBank, PhysioToolkit, and PhysioNet: Components of a new research resource for complex physiologic signals. Circulation [Online]. 101 (23), pp. e215.#x2013;e220. RRID:SCR 007345.

12. Hempel, P., Ribeiro, A., Vollmer, M., Bender, T., Dörr, M., Krefting, D., & Spicher, N. (2025). Explainable AI associates ECG aging effects with increased cardiovascular risk in a longitudinal population study. NPJ Digital Medicine, 8. 10.1038/s41746-024-01428-7.

13. Jiang, Sarah et al. Demographic reporting in biosignal datasets: a comprehensive analysis of the PhysioNet open access database The Lancet Digital Health, Volume 6, Issue 11, e871 – e878. https://www.thelancet.com/journals/landig/article/PIIS2589-7500(24)00170-5/fulltext.

14. Kittnar, O. (2023). Sex Related Differences in Electrocardiography. Physiological research, 72 Suppl 2, S127–S135. 10.33549/physiolres.934952.

15. Ko D, Rahman F, Schnabel RB, Yin X, Benjamin EJ, (2016), Christophersen IE. Atrial fibrillation in women: epidemiology, pathophysiology, presentation, and prognosis. Nat Rev Cardiol. 13(6):321–32. doi:10.1038/nrcardio.2016.45.

16. Koenig, J., & Thayer, J. (2016). Sex differences in healthy human heart rate variability: A meta-analysis. Neuroscience & Biobehavioral Reviews, 64, 288–310. 10.1016/j.neubiorev.2016.03.007.

17. Kumari, A., S., Kumar, M., Kumari, A., Kumar, S., & Dipankar, S. (2023). Effect of body mass index (BMI) on electrocardiographic P-wave dispersion among healthy adults. INDIAN JOURNAL OF PHYSIOLOGY AND ALLIED SCIENCES. 10.55184/ijpas.v75i03.197.

18. Lane, D., Skjøth, F., Lip, G., Larsen, T., & Kotecha, D. (2017). Temporal Trends in Incidence, Prevalence, and Mortality of Atrial Fibrillation in Primary Care. Journal of the American Heart Association: Cardiovascular and Cerebrovascular Disease, 6. 10.1161/jaha.116.005155.

19. Lee, K.H., Byun, S. (2023). Age Prediction in Healthy Subjects Using RR Intervals and Heart Rate Variability: A Pilot Study Based on Deep Learning. Appl. Sci., 13, 2932. 10.3390/app13052932.

20. Lin, Y., Tsai, K., Han, C., Lin, Y., Lee, J., & Lin, G. (2021). Obesity Phenotypes and Electrocardiographic Characteristics in Physically Active Males: CHIEF Study. Frontiers in Cardiovascular Medicine, 8. 10.3389/fcvm.2021.738575.

21. Ma, M., Zhi, H., Yang, S., Yu, E. Y.-W., & Wang, L. (2022). Body Mass Index and the Risk of Atrial Fibrillation: A Mendelian Randomization Study. Nutrients, 14(9), 1878. 10.3390/nu14091878.

22. Macfarlane, P. (2018). The Influence of Age and Sex on the Electrocardiogram. Advances in experimental medicine and biology, 1065, 93–106. 10.1007/978-3-319-77932-4_6.

23. Macfarlane, P.W., S.C. McLaughlin, B. Devine, T.F. Yang (1994). Effects of age, sex, and race on ECG interval measurements, Journal of Electrocardiology, 27, Supplement 1, Pages 14–19, 10.1016/S0022-0736(94)80039-1.

24. Mrad, I., Azzabi, H., Mzoughi, K., Kamoun, S., Moussa, F., Fennira, S., Zairi, I., & Kraiem, S. (2021). Correlation between body mass index and electrocardiogram parameters in healthy adults. EP Europace. 23. 10.1093/europace/euab116.015.

25. Murata, K., P.J. Landrigan, S. Araki (1992), Effects of age, heart rate, gender, tobacco and alcohol ingestion on R-R interval variability in human ECG, J. Auton. Nerv. Syst., 37, pp. 199–206, https://www.sciencedirect.com/science/article/pii/016518389290041E.

26. Ngoc, T., Nhat, L., Van, A., Thu, S., Minh, N., Tien, T., Bich, H., Viet, H., Thu, H., Phuong, N., Thuy, L., Phuong, L., Cao, C., Nguyen, L., Minh, N., Kim, C., Thu, H., & Thu, H. (2025). Electrocardiographic Profiles by sex in a cohort of healthy Vietnamese university students. American Heart Journal Plus: Cardiology Research and Practice, 60. 10.1016/j.ahjo.2025.100660.

27. Pan J. and W. J. Tompkins (1985). A real-time QRS detection algorithm, IEEE Tran. Biomedical Engineering, Vol. 32, No. 3, pp 230–236, doi:10.1109/TBME.1985.325532.

28. Pastika, L., Sau, A., Patlatzoglou, K., Sieliwonczyk, E., Ribeiro, A., McGurk, K., Khan, S., Mandic, D., Scott, W., Ware, J., Peters, N., Ribeiro, A., Kramer, D., Waks, J., & Ng, F. (2024). Artificial intelligence-enhanced electrocardiography derived body mass index as a predictor of future cardiometabolic disease. NPJ Digital Medicine, 7. 10.1038/s41746-024-01170-0.

29. Peng, X., Wang, J., Tang, C., He, L., Li, J., Xia, S., Kong, X., Zhou, N., Long, D., Sang, C., Du, X., Dong, J., &, C. (2025). Sex-specific trends in the global burden and risk factors of atrial fibrillation and flutter from 1990 to 2021. Scientific Reports, 15. 10.1038/s41598-025-93338-1.

30. Prajapati, C., Koivumäki, J., Pekkanen-Mattila, M., & Aalto-Setälä, K. (2022). Sex differences in heart: from basics to clinics. European Journal of Medical Research, 27. 10.1186/s40001-022-00880-z.

31. Quer G, Gouda P, Galarnyk M, Topol EJ, Steinhubl SR (2020) Inter- and intraindividual variability in daily resting heart rate and its associations with age, sex, sleep, BMI, and time of year: Retrospective, longitudinal cohort study of 92,457 adults. PLoS ONE 15(2): e0227709. 10.1371/journal.pone.0227709.

32. Rabkin SW, Cheng XJ, Thompson DJ. Detailed analysis of the impact of age on the QT interval. J Geriatr Cardiol. 2016 Sep;13(9):740–748. doi:10.11909/j.issn.1671-5411.2016.09.013.

33. Rao, A., Ng, A., Sy, R., Chia, K., Hansen, P., Chiha, J., Kilian, J., & Kanagaratnam, L. (2021). Electrocardiographic QRS duration is influenced by body mass index and sex. International Journal of Cardiology. Heart & Vasculature, 37. 10.1016/j.ijcha.2021.100884.

34. Rosenberg, A., Weiser-Bitoun, I., Billman, G., & Yaniv, Y. (2020). Signatures of the autonomic nervous system and the heart’s pacemaker cells in canine electrocardiograms and their applications to humans. Scientific Reports, 10. 10.1038/s41598-020-66709-z.

35. Schumann, A., & Bär, K. (2021). Autonomic Aging: A dataset to quantify changes of cardiovascular autonomic function during healthy aging (version 1.0.0). PhysioNet. RRID:SCR 007345. 10.13026/2hsy-t491.

36. Schumann, A., & Bär, K. (2022). Autonomic aging – A dataset to quantify changes of cardiovascular autonomic function during healthy aging. Scientific Data, 9(1), 95. 10.1038/s41597-022-01202-y.

37. Singleton, M., German, C., Soliman, E., Whalen, S., Bhave, P., Bertoni, A., & Yeboah, J. (2020). Body Mass Index, Sex, and Incident Atrial Fibrillation in Diabetes: The ACCORD Trial. JACC Clinical Electrophysiology, 6, 13. 10.1016/j.jacep.2020.08.008.

38. Sirichand, S., Killu, A., Padmanabhan, D., Hodge, D., Chamberlain, A., Brady, P., Kapa, S., Noseworthy, P., Packer, D., Munger, T., Gersh, B., McLeod, C., Shen, W., Cha, Y., Asirvatham, S., Friedman, P., & Mulpuru, S. (2017). Incidence of Idiopathic Ventricular Arrhythmias: A Population-Based Study. Circulation: Arrhythmia and Electrophysiology, 10, e004662. 10.1161/circep.116.004662.

39. Tan, E., J. Yap, C. Xu, Liang Feng, S. Nyunt, Rajalakshmi Santhanakrishnan, Michelle M. Y. Chan, S. Seow, C. Ching, K. Yeo, A. Richards, T. Ng, T. Lim, C. Lam (2016). Association of Age, Sex, Body Size and Ethnicity with Electrocardiographic Values in Community-based Older Asian Adults, Heart, Lung and Circulation, Volume 25, Issue 7, 705 – 711, 10.1016/j.hlc.2016.01.015.

40. Tan, S., Zhou, J., Veang, T., Lin, Q., & Liu, Q. (2025). Global, regional, and national burden of atrial fibrillation and atrial flutter from 1990 to 2021: sex differences and global burden projections to 2046—a systematic analysis of the Global Burden of Disease Study 2021. Europace, 27. 10.1093/europace/euaf027.

41. Tobenna, A. (2021). Determinants of Spatial Dispersion of P-Wave, QRS Complex, and QT-Interval on 12-Lead Electrocardiogram in Apparently Healthy Adults. Journal of Clinical Cardiology, 8. 10.23937/2378-2951/1410218.

42. Waller, B., Gering, L., Branyas, N., & Slack, J. (1993). Anatomy, histology, and pathology of the cardiac conduction system—part V. Clinical Cardiology, 16. 10.1002/clc.4960160710.

43. Wu, J., Kors, J., Rijnbeek, P., Van Herpen, G., Lu, Z., & Xu, C. (2003). Normal limits of the electrocardiogram in Chinese subjects.. International journal of cardiology, 87 1, 37–51. 10.1016/s0167-5273(02)00248-6.

44. Yang, J., Chen, Y., & Li, W. (2024). Association between body mass index and electrocardiogram indices: A Mendelian randomization study. Journal of electrocardiology, 84, 58–64. 10.1016/j.jelectrocard.2024.03.007.

45. Zheng, C., Frosted, R., Bosselmann, H., Risum, N., Joens, C., Polcwiartek, C., Christensen, H., Kragholm, K., Andersen, M., Graff, C., Torp-Pedersen, C., Bundgaard, H., & Christensen, A. (2025). The PR interval and risk of cardiac events and mortality: a nationwide study. Europace, 27. 10.1093/europace/euaf085.031.

